# Mobile instrumental assessment of swallowing in residential aged care homes

**DOI:** 10.1101/2023.07.28.23293296

**Authors:** Olga Birchall, Michelle Bennett, Nadine Lawson, Amanda Richards, Susan M. Cotton, Adam P. Vogel

## Abstract

**Purpose:** Adults living in residential-aged-care-homes (RACHs) with oropharyngeal dysphagia may not have timely access to instrumental swallowing assessment due to barriers, including the need to travel off-site for assessment. This study describes the feasibility, utility, and acceptability of mobile Flexible-Endoscopic-Swallowing-Assessment (mFEES) in Australian residential-aged-care-homes (RACHs).

**Method:** Residents with dysphagia living in RACHs were assessed using onsite mFEES. Feasibility, utility, and acceptability were measured at institutional, resident, and implementation levels.

**Result:** Healthcare professionals and medical decision makers reported that mFEES facilitated a better understanding of residents’ swallowing function/dysphagia management and was beneficial over off-site services. Self-rated discomfort during mFEES was low and most residents presented with no or minimal anxiety about the procedure. Costs of mobile assessments are documented.

**Conclusion:** mFEES was a safe, well tolerated, and practical service that offered opportunity to enhance person-centered clinical care in older adults living with dysphagia in RACHs.

## Introduction

Many older adults living in residential aged care homes (RACH) experience swallowing difficulties-oropharyngeal dysphagia (OD), as a result of physiological or anatomical changes associated with ageing, frailty and/or coexisting medical conditions (e.g., dementia). OD is a geriatric syndrome with prevalence rates in the RACH setting ranging between 40 and 68% (Baijens et al., 2016; Nogueira & Reis, 2013; Steele, Greenwood, Ens, Robertson, & Seidman Carlson, 1997).

OD can lead to serious complications, including dehydration, malnutrition, nutrition-related sarcopenia (Wakabayashi, 2014), aspiration (entry of food, fluids, or secretions into the airway), choking and reduced quality of life. In a RACH setting, mortality rates for adults with dysphagia have been reported to be higher than for adults without dysphagia (Jukic Peladic et al., 2018) and choking has been described as the second leading cause of preventable deaths (Ibrahim et al., 2017).

Timely diagnosis and effective dysphagia management can optimise the quality of care and swallowing related outcomes while reducing the incidence of dysphagia related complications (Rosenvinge & Starke, 2005). Yet, the quality of dysphagia diagnosis and management in the RACH setting has been questioned with calls for early dysphagia diagnosis and person-centered, evidence-based interventions (Chen, Kent, & Cui, 2021; Jukic Peladic et al., 2018).

Flexible endoscopic evaluation of swallowing (FEES) is an evidence-based instrumental swallowing assessment tool commonly used outside of the RACH setting in adults with dysphagia to supplement the findings of a clinical swallowing examination (CSE). FEES involves passing a nasendoscope with a light source and a camera (at its tip or attached to a camera head) transnasally to visualise structures of the pharynx and larynx, secretion management, and passage of food/drinks while swallowing (S. E. Langmore et al., 2022). FEES allows healthcare professionals involved in dysphagia care, including speech-language-pathologists (SLP), to establish a more specific dysphagia diagnosis. This information can be used to tailor dysphagia treatments and education.

Emerging evidence suggests that despite its clinical utility, potential to support informed decision making and education, FEES is not commonly used in adults in RACHs (Birchall, Bennett, Lawson, Cotton, & Vogel, 2021a, 2021b, 2022; Rogus Pulia, Wirth, & Sloane, 2018). There are barriers that might limit the use of FEES in a RACH setting (Birchall et al., 2021b; Rogus Pulia et al., 2018). Many of these relate to the need to travel to an outpatient clinic in order to access FEES, introducing: travel associated costs; difficulty organising accompanying staff; as well as the physical burden of transportation to residents with frailty, increased risk of falls, dementia, and anxiety (Birchall et al., 2021b). Access barriers may be amplified for residents living in geographically remote locations with fewer specialist dysphagia clinics and greater distances between the RACHs and outpatient clinics offering ISA. Access may also be reduced during infectious disease outbreaks (e.g., COVID-19) due to: legislated travel restrictions, personal preference of residents who are reluctant to leave their homes and risk disease exposure (Mehrotra, Chernew, Linetsky, Hatch, & Cutler, 2020), and because hospitals may be less likely to offer timely outpatient appointments (Mehrotra et al., 2020; Muschol & Gissel, 2021; Prvu Bettger et al., 2020).

In the US and Japan, mobile FEES (mFEES) is offered onsite in some RACHs, with authors suggesting potential benefits to this service model (Barczi, Sullivan, & Robbins, 2000; Birchall et al., 2021a; Hase et al., 2019; Takahashi, Kikutani, Tamura, Groher, & Kuboki, 2012). A recent survey of SLPs in Australia, where onsite FEES is currently unavailable, revealed professional support for mFEES to be trialled in RACHs (Birchall et al., 2021b). This finding is supported by recent updates to SLP professional body guidelines in Australia and the UK to allow the provision of mFEES in a RACH setting (Australia, 2019; Kelly et al., 2015).

To date, however, there have been no studies evaluating mFEES as a service in Australian RACHs, nor studies considering the multiple stakeholders and potential levels of implementation. Therefore, it is not known how a mFEES service impacts: (i) the resident, (ii) RACH staff and the RACH as an institution, and (iii) to what extent mFEES is practical to conduct in a RACH setting and will meet the residents’ healthcare goals (implementation level). Thus, the aim of the study was to describe the feasibility of a mFEES model in RACHs including practicability, utility (ability to meet healthcare goals), and acceptability (safety and tolerability) of mFEES.

## Materials and Methods

### Study Design

A feasibility study of onsite ISA with 12 residents living in RACHs in metropolitan and regional Victoria, Australia was conducted between February and October 2021.

### Sample and Setting

Convenience sampling was used to select RACHs. Facilities were recruited if they were: (i) located within 3 hours drive from metropolitan Melbourne, Australia; (ii) accredited by the Australian Aged Care Quality Agency; and were able to (iii) provide a specified selection of foods/fluids and utensils for the swallowing assessment (Appendix 1); (iv) allocate a division 1 nurse to be present during the mFEES procedure to provide clinical monitoring of the resident; (v) comply with the studies’ COVID-19 safety protocol (Appendix 2); and (vi) demonstrate that existing SLP services were happy to support the mFEES study.

Residents were recruited from participating RACHs if they were identified by their SLP as having potential to benefit from an ISA, met mFEES inclusion criteria developed for this study and based on based on Langmore and Aviv (2001) (S. Langmore & Aviv, 2001) (Appendix 3).

### Measures

Measures of study feasibility, utility and acceptability were collected across the RACHs, residents and implementation levels (Table 1).

**Table 1.** Measure of mFEES study feasibility, utility and acceptability across RACH, participant, and implementation levels.

| Areas of consideration | Level of analysis | Measures Used |
| --- | --- | --- |
| **Feasibility** | RACH | • division 1 nursing time per mFEES<br>• cost of personal protective equipment (PPE) supplied by the RACH per mFEES (cost estimates in appendix-average cost per patient) |
|  | Resident | • referral response rate (days) |
|  | Implementation | • referral rate<br>• percentage of appropriate referrals<br>• percentage of eligible residents referred by their treating SLP who agreed to the mFEES<br>• percentage of successful mFEES studies completed (i.e., nasendoscope successfully inserted and procedure terminated based on the clinical needs of a resident)<br>• SLP time in minutes (i.e., travel time, time onsite at the RACH, mFEES report preparation time)<br>• cost of individual mFEES assessment (i.e., mFEES equipment/perishables, personal protective equipment)<br>• considerations in the physical set-up of the procedural space for mFEES |
| **Utility** | RACH | • post-mFEES survey |
|  | Resident | • mFEES report<br>(i) did the study answer clinical questions posed by the referring SLP,<br>(ii) did the information and images included in the report provide opportunities for education of residents, and families |
| **Acceptability** | Resident | • mFEES duration<br>• pre-procedural anxiety (visual facial anxiety scale for assessing preoperative anxiety[13]), procedural discomfort (Faces Pain Scale – Revised [14, 15]),<br>• number and nature of procedural mFEES adverse events |

In addition, since mFEES was part of clinical care, for each participant the following information was collected: basic biographic details (e.g., age, medical diagnoses, current food, and fluids), oral hygiene status (i.e., as measured by the Oral Health Assessment Tool, OHAT (Chalmers & Johnson, 2004), and mFEES assessment findings. Measures used as part of the mFEES report included the Marionjoy Secretion Rating Scale (Donzelli, Brady, Wesling, & Craney, 2003) (to measure pharyngeal and endolaryngeal secretions), the Penetration-Aspiration Scale (PAS) (Rosenbek, Robbins, Roecker, Coyle, & Wood, 1996) to measure airway protection from oral intake; the Yale Pharyngeal Residue Severity Rating Scale (Leder & Neubauer, 2016) to measure pharyngeal food/fluid residue; and the Functional Oral Intake Scale (FOIS) (Crary, Mann, & Groher, 2005) to rate the overall level of oral intake.

### Procedures

Ethical approval was granted by The University of Melbourne Human Research and Ethics Committee (reference number 021-13387-13943-2, 08/02/21). An electronic advertisement for the study with links to the plain language statement and consent form was posted in online professional SLP groups, emailed to RACH facility managers, private SLP service providers and SLP hospital outpatient clinics. Facilities were recruited in chronological order of consent if they satisfied the study inclusion criteria, signed the study consent form, and had a private SLP who was supportive of the study.

SLP servicing RACHs recruited into the study were provided with mFEES referral criteria and the researcher’s contact details to discuss potential referrals. Residents of participating RACHs who met these inclusion criteria were referred into the study by their treating SLP. Referrals were also conditional on support from the treating general practitioner (GP) and/or geriatrician.

Written consent was sought from the residents and/or their legally appointed medical decision makers (MDMs), and all individuals present during the mFEES (i.e., MDMs, nursing staff, treating SLP).

Consultations occurred in the residents’ bedrooms with doors closed to optimise privacy, comfort, and aerosol containment. Where practical, windows were opened to increase ventilation.

The research-SLP (R-SLP) screened oral hygiene and performed a limited cranial nerve examination prior to the endoscopy. Oral hygiene and cranial nerve screening are common components of SLP-led swallowing examinations because cranial nerves control the muscles involved in swallowing, while compromised oral health can affect swallowing function and is a recognised risk factor for aspiration pneumonia, in adults with OD (van der Maarel-Wierink, Vanobbergen, Bronkhorst, Schols, & de Baat, 2011).

Residents rated their pre-procedural anxiety using the Visual Facial Anxiety Scale (VFAS) (Cao et al., 2017).

The R-SLP set-up mFEES equipment, food/ fluids to be trialled, and consumables (Appendix 4) in the residents’ bedrooms, ensuring easy access to a sink (for hand hygiene during mFEES equipment high-level disinfection), emergency alarm and a rubbish bin. Reusable and a disposable rhino-laryngoscopes were used to conduct the mFEES. High level disinfection of the reusable rhino-laryngoscope was conducted using the Tristel Trio^TM^ Wipes System (Tristel Solutions Ltd, Cambs., UK) immediately before and after use. To minimise the hypothetical risk of COVID-19 transmission, all family/staff present in the room during the mFEES wore high-level PPE.

The R-SLP coordinated the mFEES, collected supplementary assessment data (i.e., oral hygiene, bulbar screening, measures of procedural anxiety and discomfort), passed the nasendoscope, monitored endoscopy tolerance and provided instructions/reassurance to the resident (shown in Fig. 1). The nurse and/or referring SLP (if present) provided residents with food/fluids, and operated mFEES recording equipment, as instructed by the R-SLP during the mFEES. They also assisted residents to maintain appropriate positioning through gentle tactile and verbal cues and offered additional reassurance, when indicated. The mFEES adverse event protocol (Appendix 5) was available to support clinical decision making and to manage clinical risks. Nursing staff were responsible for implementing facility protocols in the event of medical complications. The R-SLP determined the order, volume and type of oral intake trialled based on: (i) information provided by the referring SLP (e.g., clinical swallow assessment findings, goals of the mFEES including the resident’s wishes); (ii) R-SLP’s clinical observations/ reasoning during the study; and (iii) mFEES procedural guidelines (S. E. Langmore et al., 2022). Residents, nursing staff, SLP and legally appointed MDMs could watch the mFEES with the R-SLP on a portable screen during and immediately after the study.

**Fig. 1.**
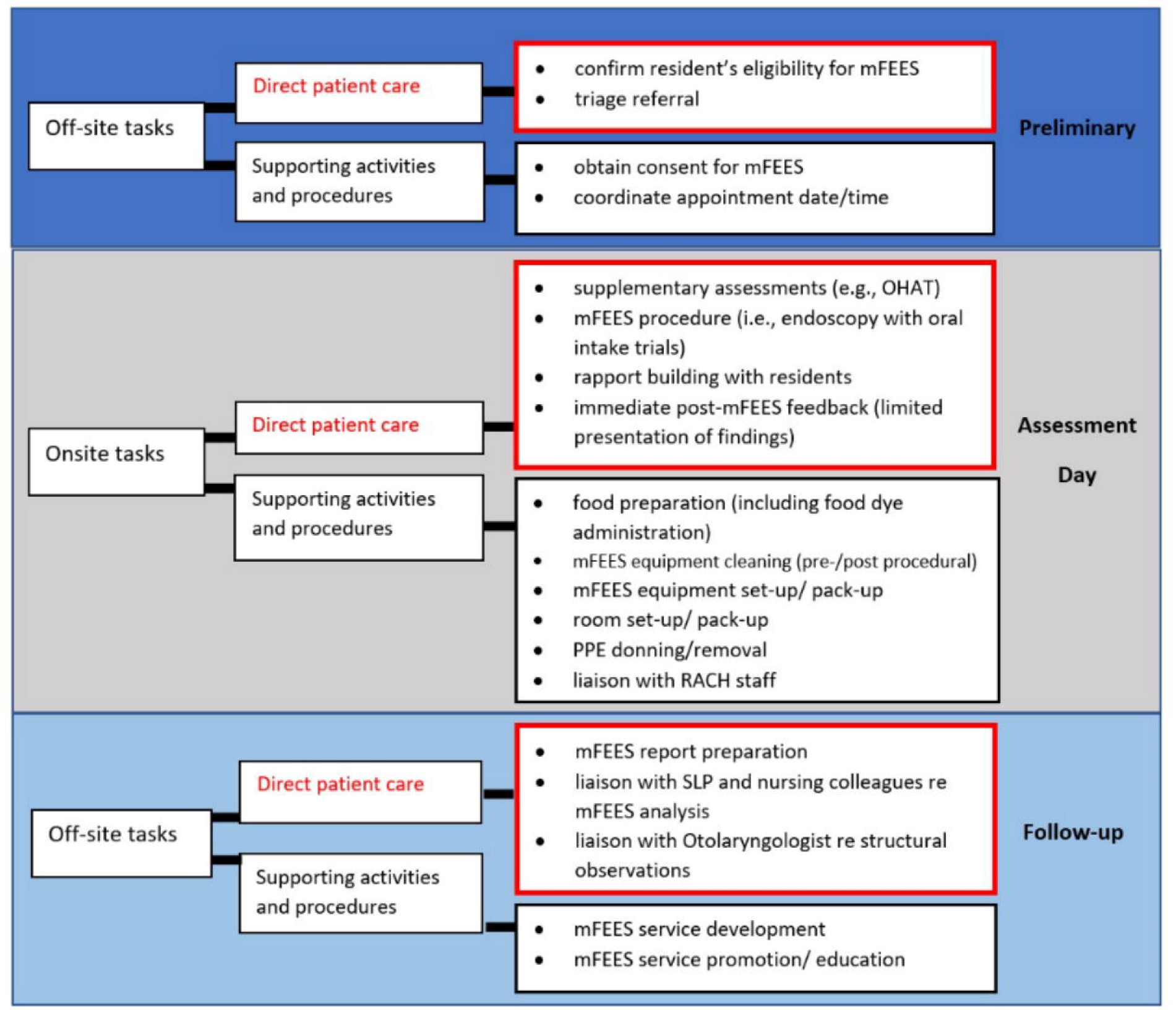
Speech Language Pathologist’s responsibilities in providing mFEES to adults in RACHs.

The R-SLP and another experienced SLP (NL) viewed, discussed, and analysed mFEES recordings in real-time and frame-by-frame using the VLC Media Player (3.0.14) software on a Microsoft Surface Laptop (model 1769). An Otolaryngologist experienced in FEES and swallowing (AR) viewed and commented on anatomical observations, when requested by the R-SLP. The R-SLP prepared a mFEES report using a mFEES report proforma (Appendix 6). A copy of the report was sent via email to the referring SLP, usually within one week of the assessment. The treating SLP was responsible for ongoing dysphagia care (including communication of mFEES findings to residents, healthcare staff and families).

Nursing staff, SLP and MDMs completed a post-mFEES survey containing 12 questions about their experience with the mFEES service (Appendix 7). Most were closed questions (e.g., ‘Did the mFEES provide you with useful information about the participant’s swallowing function?’), with multiple choice response options (i.e., ‘Yes’, ‘No’ or ‘undecided’). There were two questions requiring participants to rate their experience on a ten-point Likert scale (e.g., *On a scale of 1-10, where 1 is completely unsatisfied and 10 is completely satisfied, how would you rate your satisfaction with this mobile FEES experience*?). For each question participants could make qualitative comments about their response. Data were collected and managed using REDCap electronic data capture tools hosted at the University of Melbourne (Harris et al., 2019; Harris et al., 2009).

### Statistical Analyses

Quantitative data were analysed in IBM® SPSS®, Version 26.0 (2019. Armonk, NY: IBM Corp). Demographic characteristics and survey responses were summarised using descriptive statistics. Agreement in response to survey questions was defined as 70% or above consensus about an issue. This criterion has been commonly used in healthcare research and is believed to represent clinically meaningful and reproducible consensus (Birchall et al., 2021b; Gephart, Effken, McGrath, & Reed, 2013)

## Results

The study occurred over an 8-month period (01/03/21-29/10/21) to ensure completion within the research funding period. Seventeen referrals were initiated by SLP during this time. Of eligible residents referred into the study, 3 referrals were withdrawn before consent was sought due to: deterioration in medical status (1 case), RACH internal staffing changes (1 case), lack of support from the medical practitioner (i.e., GP preferred a different ISA). Of the remaining 14 referrals, written informed consent was provided by 85.7% (*n*=12, *N*=14) of residents. Participant demographics are captured in Table 2.

**Table 2.** Participant demographics.

| Characteristics |  | Percentage of residents (*n*) |  |
| --- | --- | --- | --- |
| Male gender |  | 50 | (6) |
| Locality | Regional | 66.7 | (8) |
|  | Metropolitan | 33.3 | (4) |
| Four or more medical comorbidities |  | 91.6 | (11) |
| Cognitive impairment (i.e., due to dementia, acquired brain injury, PD, stroke) |  | 58.3 | (7) |
| Parkinson's disease (PD) |  | 33.3 | (4) |
| Oesophageal dysphagia (i.e., GORD, Schatzki's ring, severe oesophageal dysmotility) |  | 66.7 | (8) |
| Diet<sup>a</sup> | Regular | 25 | (3) |
|  | Soft and bite sized | 25 | (3) |
|  | Minced and moist | 33.3 | (4) |
|  | Pureed | 16.7 | (2) |
| Fluids<sup>a</sup> | Regular | 41.7 | (5) |
|  | Mildly thick | 41.7 | (5) |
|  | Moderately thick | 16.7 | (2) |
| OHAT score | 0-2 | 33.3 | (4) |
|  | 3-5 | 58.3 | (7) |
|  | > 6 | 8.3 | (1) |

<sup>a</sup>Consistencies were not mutually exclusive.

### Practical Considerations in Providing mFEES Services

#### Referral Response Rate

In considering practical aspects of conducting the mFEES service at the level of the resident, we note that on average mFEES assessments occurred within 7.6 days of referral. Referral response was affected by Victorian government COVID-19 related regulations, including episodic restrictions on entry into RACHs and travel between metropolitan and regional Victoria.

#### mFEES Service Provider Resources

The R-SLP travelled up to 4 hours (return trip) per referral. The average total time onsite at the RACH was 88.7 minutes (m) (range 45-120m). Direct resident contact time was 31.7m on average (range 25-51.7m), including endoscopy (average 16.7m, range 11.7-21.9m). The R-SLP prepared mFEES reports within 45 minutes, including interdisciplinary liaison time. Otolaryngology support was sought on three occasions (15 minutes each) to comment on anatomical observations.

The capital cost of re-usable mFEES equipment was AUD$59,954.00, while disposable set-up was only AUD$7,508.00 (Table 3). The average per capita cost of conducting a mFEES was $Au179.97 using re-usable and AUD$338.00 using disposable equipment set-up (Table 4).

**Table 3.** Capital costs of mFEES equipment.

| **Reusable set-up**<br>(Karl Storz products) |  | **Disposable set-up**<br>(Ambu® products) |  |
| --- | --- | --- | --- |
| **Item** | **Cost**<br>**(AUD\$)** | **Item** | **Cost**<br>**(AUD\$)** |
| Strobo-Video-Rhino-Laryngoscope (TS5854) in transportation case (TP – 100 Tele Pack x LED) | 38,000.00 | n/a | n/a |
| Telepack X ED with silicone keyboard (20040240US) and leak testing manometer | 21,946.00 | Ambu® a View™ 2 Advance portable monitor | 7,500.00 |
| Toshiba 32 GB USB 2.0 Flash Drive | 8.00 | Toshiba 32 GB USB 2.0 Flash Drive | 8.00 |
| **Total** | **59,954.00** |  | **7,508.00** |

**Table 4.**
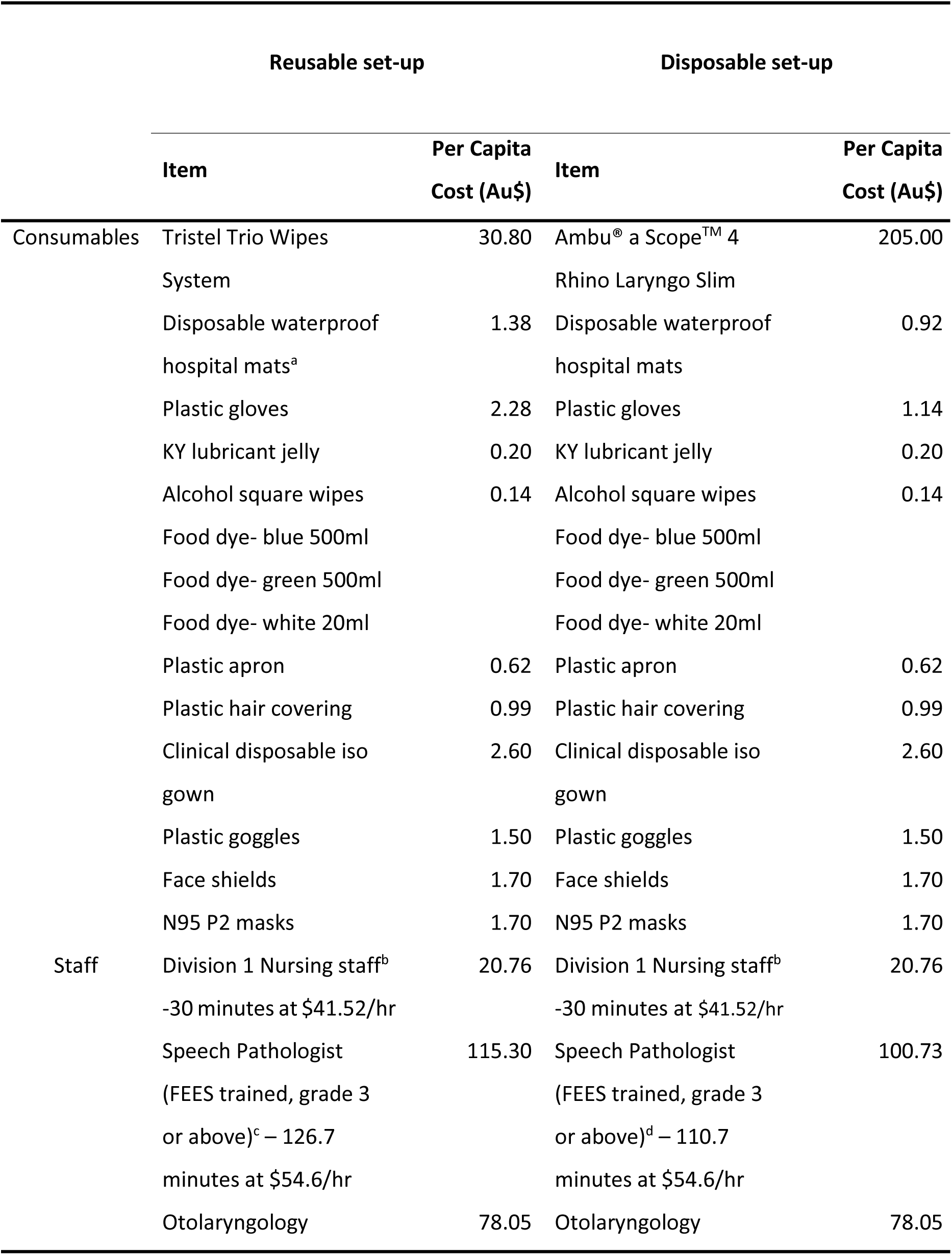
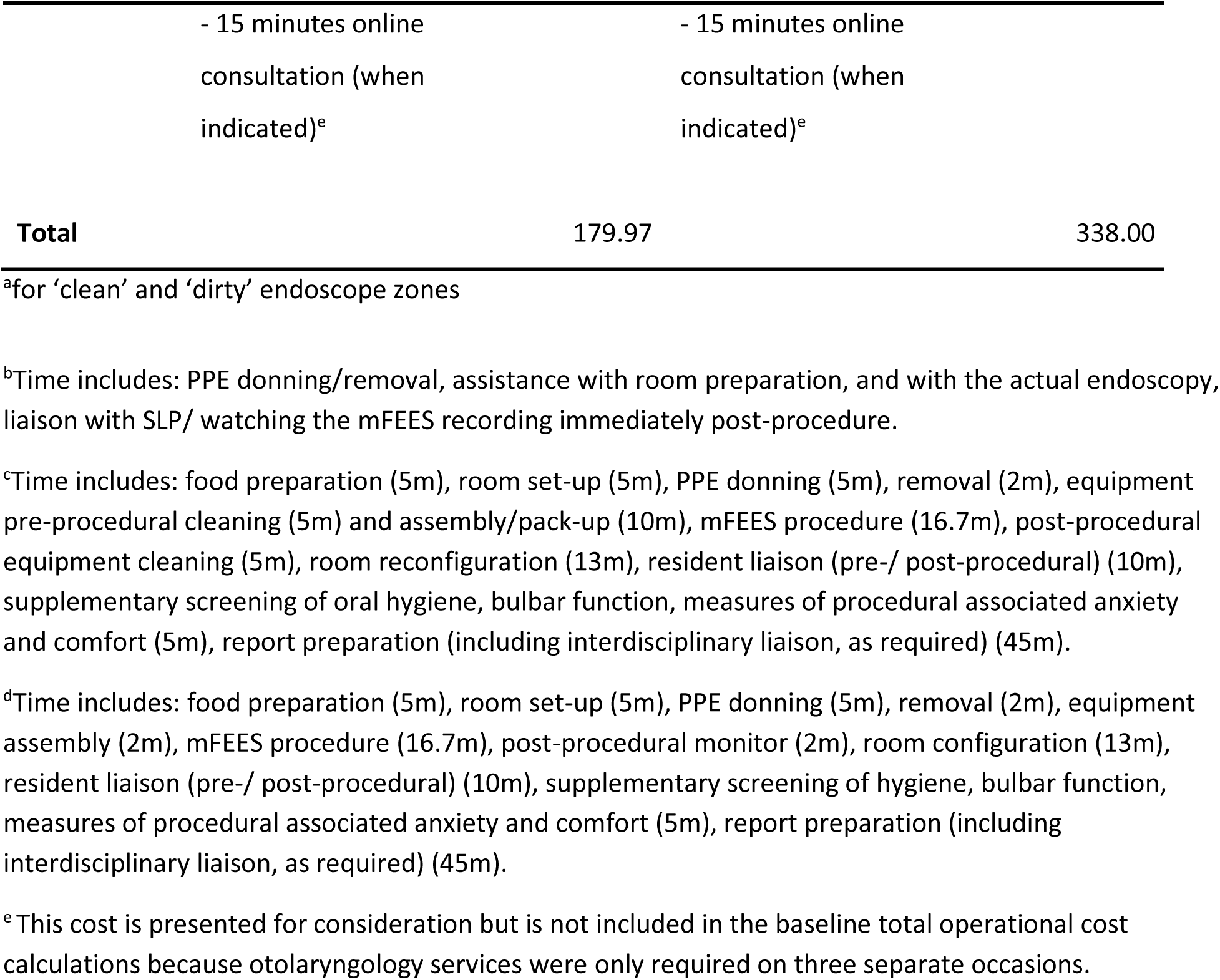
Operational costs of mFEES equipment.

#### Procedural Space

Practical considerations identified in setting up bedrooms for mFEES included: (i) size (allowing unobstructed movement of up to 4 adults), (ii) ventilation (for thermoregulation and clearance of circulating fumes from disinfection products), and (iii) availability of flat and stable surfaces(shown in Fig. 2). to accommodate the: (i) mFEES screen; (ii) nasendoscope, lubricant and alcohol wipes; and (iii) food/fluids.

**Fig. 2.**
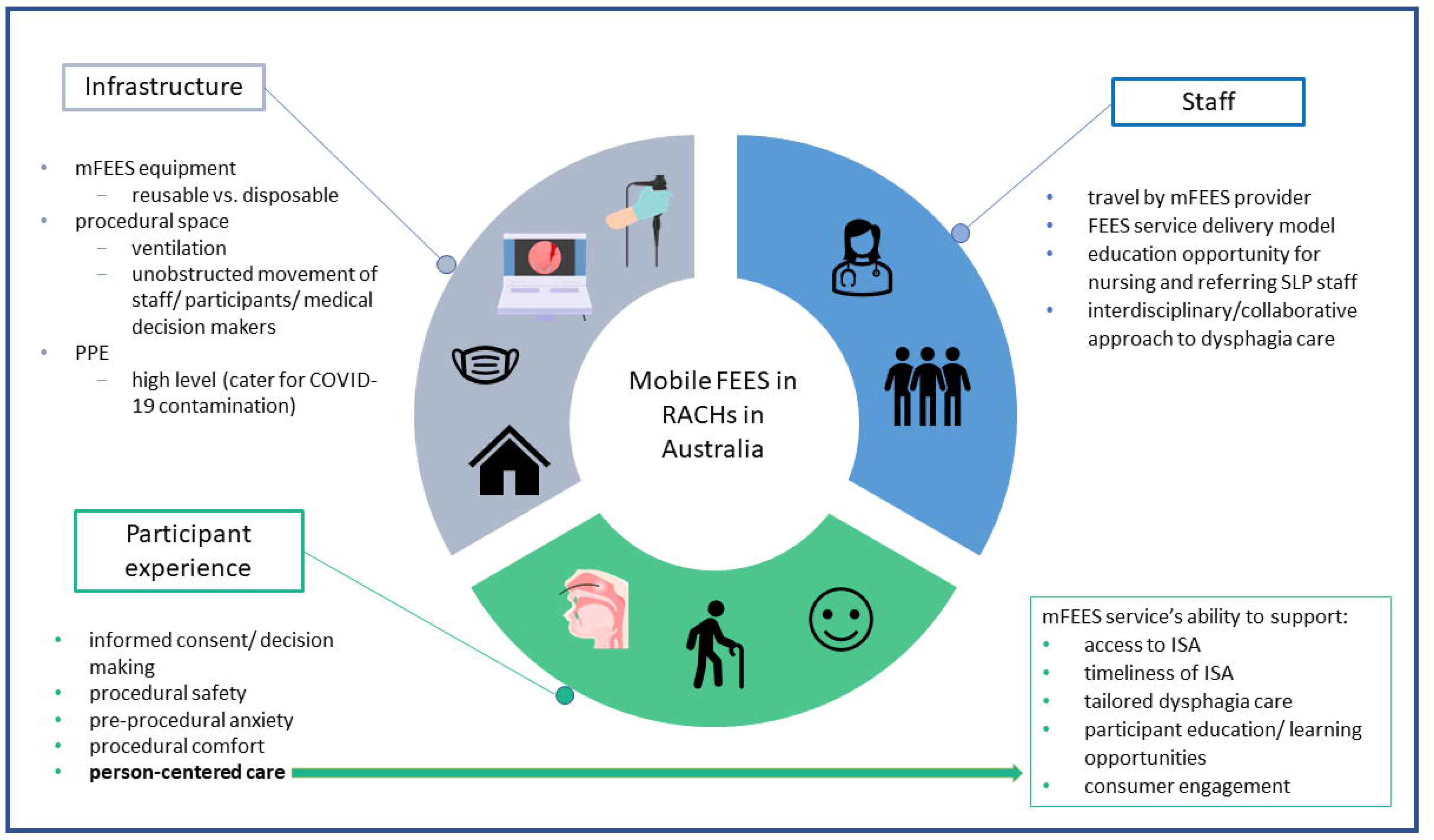
Considerations in providing mFEES services in RACHs in Australia.

#### RACH Resources

The mFEES service utilised facility resources. RACHs nursing staff were present for an average of 30 minutes per mFEES study (range 15-45m), including donning of PPE, procedural aspects of mFEES, liaison with SLP, and optional viewing of the mFEES video. Beyond contributing nursing time, RACHs supplied standard PPE for up to 3 attendees (i.e., facility SLP, nursing staff and MDMs). The cost of PPE per mFEES for the RACH was $8.07 per person ($24.21 for 3 attendees). RACH staff and families believed that the mFEES referral was easy to organise (92.6%, *n*=25, *N*=27) and were satisfied with referral response rates (96.3%, *n*=26, *N*=27). Staff commented on the “excellent turnaround time between referral and mFEES assessment”, but acknowledged that “organisational barriers”, “COVID-19 lockdowns”, and the need for “consent from multiple stakeholders” slowed referrals at the RACH level.

### Utility of mFEES Services

#### Opinions of RACH Nursing Staff, Referring SLPs, and Medical Decision Makers

Opinions of healthcare professionals and MDMs on the utility of mFEES are summarised in table 5.

**Table 5.** Opinions of healthcare professionals and medical decision makers on the utility of mFEES services.

| Perceived benefit of mFEES | Percentage of<br>survey respondents<br>who agreed (n/N) |  | Respondent's comments (examples) |
| --- | --- | --- | --- |
| Provides useful information | 100 | (26 / 26) | "being able to..visualise [anatomy] during aspiration and attempted clearing coughs was valuable for all..."<br>"allowed me to balance [QoL] with risk much more accurately"<br>"images to show the [resident] when providing Dignity of Risk education"<br>"gives space for providing recommendations more holistically because usually we need to be extremely cautious around any overt signs of penetration/aspiration"<br>"added significant weight to working diagnosis"<br>"..provided information about clinical risk..."<br>"helped with clinical decision making..evidence to support my recommendations" |
| Increased staff/family/resident's understanding of: |  |  |  |
| swallowing function | 96.2 | (25/26) | "RACH staff are often time poor and [without mFEES] may not have any understanding of silent aspiration or the impacts of dysphagia" |
| dysphagia management | 84.6 | (22/26) |  |
| Assessment was conducted: |  |  |  |
| in a familiar environment | 100 | (26/26) | "..it would have been very stressful for the resident if she was to travel to an unfamiliar place.."<br>"..residents ..feel comfortable and safe in their own environment.." |
| with familiar staff/ family present | 74.1 | (20/27) | "RACH staff are often time poor and may not [understand] silent aspiration or the impact of dysphagia"<br>"..difficult and lengthy process to educate family/ staff about benefits of a FEES to the point where they..agree to travel.." |
| at an appropriate time for the resident | 70.4 | (19/27) | "enabled resident involvement" |
| High satisfaction with service model (rating 8-10<sup>a</sup>) | 100 | (26/26) | "so happy that the service came to my dad..so he wasn't caused any more distress"<br>"easy to organise"<br>"competent and caring ..[speech pathologist] conducting the FEES"<br>"accuracy and good knowledge"<br>"...responsive service that facilitated health promotion.."<br>"...gold standard [swallow assessment] in the person's room"<br>"brilliant, patient focused care" |
| Mobile service model is useful compared to an off-site model (rating 9-10<sup>b</sup>) | 100 | (26/26) | "most of the residents I work with have....factors [that] make it unfeasible to travel"<br>"without mFEES..[resident] would ...[need] to travel 60 minutes... which was a barrier"<br>"...[increased] accessibility of the ISA.."<br>"able to visit people who are unable to attend major hospitals"<br>"quick access to instrumental assessment in resident's environment"<br>"..allows timely discussion of results/management"<br>"..other staff members were also able to gain insight into swallowing" |
| Mobile FEES should become a standard optional part of swallowing assessment and management in adults living in RACHs | 92.0 | (23/25) | "..help [SLP] to make more appropriate dysphagia management recommendations"<br>"..empower our residents to take control of their dysphagia.."<br>"there are residents who could implement strategies successfully instead[of modified diet/fluids] or who are not as 'at risk' as we believe they are" |

<sup>a</sup> satisfaction rating where 1=completely unsatisfied and 10=completely satisfied
<sup>b</sup> usefulness rating where 1=not useful and 10=highly useful

#### mFEES Reports

At the resident level, in all cases the mFEES report addressed clinical questions documented by the referring SLP in the mFEES referral form (Table 6). Information derived through mFEES could be used to tailor dysphagia care and to educate residents, their families, and staff (Appendix 8). For example, a resident requesting to eat toast demonstrated silent aspiration of solid food during the mFEES. Images of this event were incorporated into the mFEES report. The treating SLP could use these images to educate the resident about his or her personal risks of eating toast to facilitate informed decision making. In another case, the mFEES demonstrated aspiration of oral intake and frothy secretions that returned through the cricopharyngeal sphincter after completion of the swallow (information that is impossible to obtain with CSE alone), providing an opportunity for the medical team to review the effectiveness of existing GORD management strategies. Another resident receiving texture-modified oral intake reported waiting seven months for a repeat ISA. mFEES findings revealed that compensation with small mouthfuls reduced silent aspiration and pharyngeal residue more effectively than increased fluid viscosity (thickened fluids) enabling the SLP to provide more tailored compensatory dysphagia strategies.

**Table 6.**
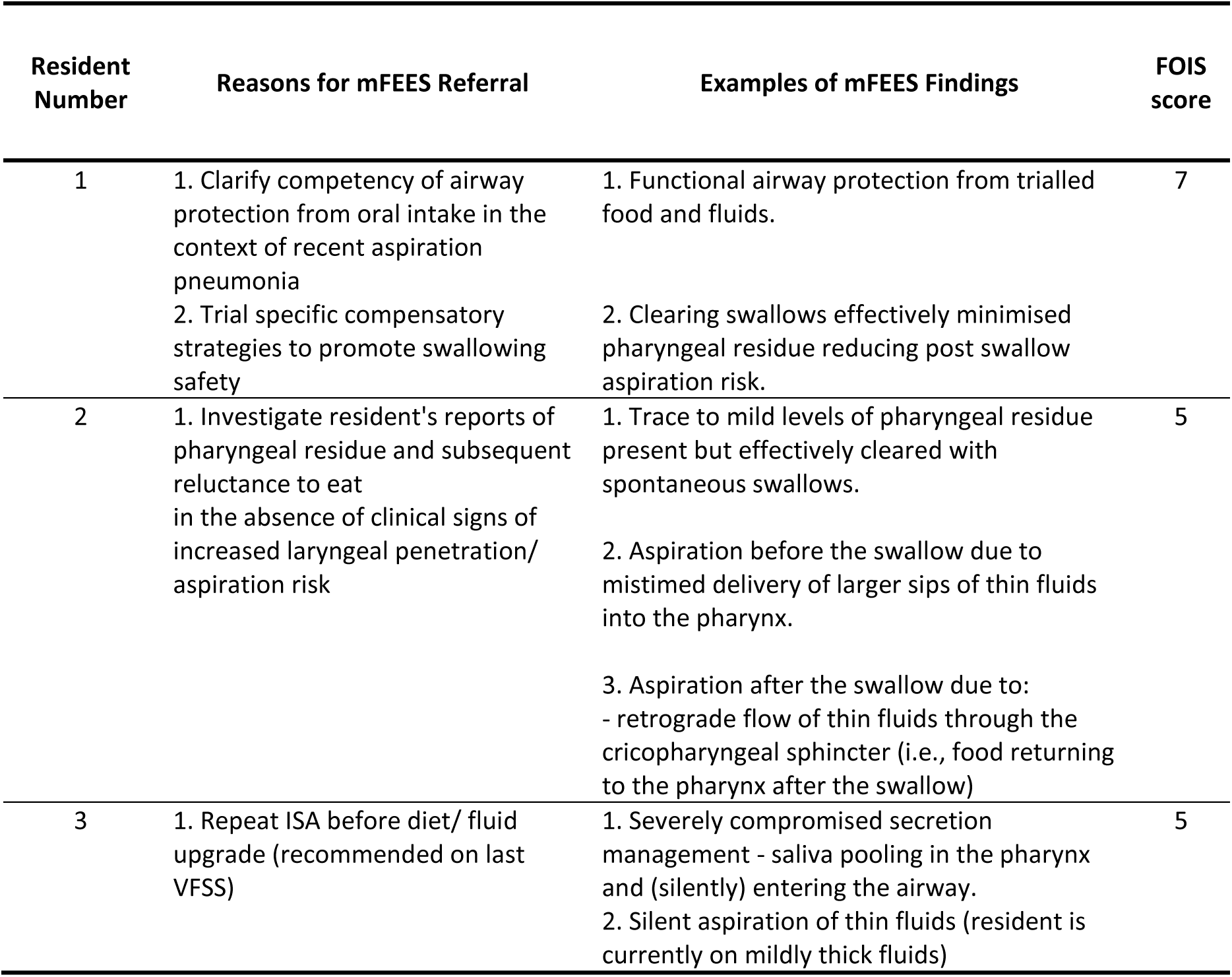
Examples of reasons for mFEES referral and key mFEES findings.

### Acceptability of mFEES Services

The safety and tolerability of the onsite mFEES service model was explored at RACH and resident levels.

#### mFEES Safety

There were no instances of the mFEES being ceased due to safety concerns.

#### mFEES Tolerability

In the post-mFEES survey, five respondents (18.5%; *n*=5, *N*=27) identified nasendoscope tolerance as an area of consideration. Respondents commented on residents with cognitive impairments experiencing heightened procedural anxiety, potential discomfort and difficulty avoiding extraneous movements with the nasendoscope in-situ.

Fifty percent of residents rated their level of pre-procedural anxiety as ‘none’ or ‘mild’. Two residents reported their level of anxiety to be ‘mild-moderate’, ‘moderate’, and ‘moderate-to-high’, respectively. Residents with the highest anxiety ratings, had reports of pre-existing anxiety documented in their medical histories or reported by NS. Average self-rated discomfort during the mFEES was 2.83 (range 0-6) on a 10-point rating scale with 2-point selection increments. There was one instance of a very mild, self-limiting nosebleed in a resident with intellectual disability who presented with moderately dehydrated and crusty nasal mucosa. A sudden head movement during nasendoscope insertion caused dislodgement of a small mucosal crust. The nasendoscope was withdrawn to ensure resident comfort. Gentle nasal hygiene was performed with warm, moist cotton swabs to loosen remaining crusty secretions, and the nasendoscope was reinserted through the same nostril with the resident’s consent without difficulty or reported discomfort.

## Discussion

This study trialed and demonstrated the feasibility, utility, and acceptability of a SLP-led mFEES service model for older adults living in Australian RACHs. After considering this model at a resident, RACH, and implementation level, we suggest that mFEES in a RACH setting is a safe, well tolerated, and practical service that offers opportunities to enhance clinical care of older adults with dysphagia.

Moving forward, clinical guidelines stipulating appropriate mFEES referral criteria, as well as minimal infrastructure, governance, and fiscal requirements would help providers to establish a viable mFEES service.

### Practical Considerations in Providing mFEES Services in RACHs

#### Recruitment of Participants

This study was challenged by the: (i) COVID-19 pandemic, and (ii) novelty of the service model. This was the first time that mFEES services have been delivered in Australian RACHs. Hence, RACHs took time to develop clinical governance process to support onsite endoscopy. Some RACH managers/nursing staff reported being unfamiliar with FEES, its role, risks, and potential benefits. This may be because off-site FEES is rarely performed with adults living in RACHs due to access barriers (e.g., the need to travel off-site for assessment) (Birchall et al., 2021b). Most study participants resided in regional Victoria, where access barriers may be amplified compared to metropolitan Melbourne, due to longer distances between healthcare providers and potentially a smaller number of SLP clinics with specialised FEES training/equipment. One participant in our study reported waiting seven months to access an ISA. Recruitment was also prolonged by unprecedented state government, and local facility restrictions on entry into RACHs and healthcare provision (particularly aerosol generating procedures) in response to the COVID-19 pandemic. There were periods when the R-SLP was unable to travel between metropolitan and regional facilities and/or unable to perform FEES, which was considered to be an aerosol generating procedure. Referring SLPs also reported a decrease in referrals by the RACHs, resulting in a smaller pool of potential mFEES referrals.

#### Infrastructure Considerations in Providing mFEES

Our model trialed both re-usable and disposable mFEES equipment. Compared to the disposable unit, re-usable equipment provided perceptually superior image quality, and captured clear sound in-line with the video recordings. However, the equipment was larger (required more space upon assembly), heavier, and more cumbersome to manoeuvre; took slightly longer to assemble; had to be plugged into a power point during usage; and the nasendoscope required high-level disinfection between uses. The R-SLP prioritised superior image quality and hence reusable equipment was used for the majority of procedures in this study. However, the disposable unit provided a viable alternative in the event of technical malfunction and in cases where other aspects of clinical care required priority (e.g., insufficient space/time for Karl Storz equipment cleaning/assembly).

In our study, bedrooms were chosen to provide mFEES because they offered residents the comfort of a familiar environment, privacy, relative containment of aerosols (with the door closed), access to a sink, and a bed (to enable postural adjustment in the unlikely event of a vasovagal episode).

#### RACH Nursing Staff Availability to Assist in mFEES

There are international shortages in resident-to-nursing ratios, which have been exacerbated by the COVID-19 pandemic (Australian Nursing & Midwifery Federation, 2020; Grabowski & Mor, 2020; Xu, Intrator, & Bowblis, 2020). In Australia, over 50% of residents live in RACHs with unacceptable staffing levels (Eagar K & Centre for Health Service Development, 2019), meaning that RACHs may lack the resources to allocate support workers to travel off-site with vulnerable residents for ISA. Our mFEES model reduced demands on nursing time by: eliminating travel; requiring nursing attendance only during the nasendoscopy; and where possible scheduling assessments during overlapping morning-afternoon shifts.

#### mFEES Service Model

This study utilised the cheapest FEES service delivery model where one SLP and one RACH nurse were the only essential personnel, and high-level disinfection (rather than sterilisation) was used to clean the nasendoscope (Cimoli & Sweeney, 2012). This model was predicated on the R-SLP having expertise in all aspects of FEES (endoscopy and FEES analysis) and sufficient clinical experience to direct supporting staff (e.g., in presenting appropriate food/fluids and equipment operation) while performing the nasendoscopy. The R-SLP performed allFEES clinic set-up, implementation, and cleaning activities. Published data on the total duration of these activities is limited. One published cost analysis suggests that 30 minutes of direct patient contact time is required for the SLP and the nursing staff to conduct the procedure (i.e., cumulative time = 60 minutes) and 20 minutes for one person to perform high level disinfection (Cimoli & Sweeney, 2012). Our findings align with these data. The R-SLP also traveled 0.5-4 hours (return) per resident to provide assessment. Extended travel was possible for the purposes of the current study, yet the cost of travel and the number of residents seen within a geographical location are important considerations in establishing future larger scale, financially viable mFEES services.

### Utility of mFEES Services in RACHs (clinical risk and education)

The mFEES service enabled residents to receive timely, person-centered care-in-place while minimising COVID-19 exposure risks associated with travel to an outpatient hospital clinic. Since many hospital-based clinics operated with reduced capacity during the COVID-19 pandemic (Alfred Health, 2021), our model offered older adults access to ISA services that may not have been available to them during the upsurge in COVID-19 cases.

#### Tailored Dysphagia Care

Information obtained during the mFEES assessments addressed clinical questions posed by the referring SLPs including queries around secretion management, pharyngeal/laryngeal anatomy, and swallowing dynamics with clinical findings to formulate increasingly: differential diagnoses, tailored swallowing management and targeted education. In this way the clinical risk of complications from dysphagia and inappropriate dysphagia management could be reduced. For example, in RACH thickened fluids are commonly provided to adults identified to be at risk of aspirating regular fluids based on CSE findings. Iatrogenic complications of thickened fluids include dehydration (Barker, Craig, Spiers, Kunonga, & Hanratty, 2018) (one of the top four causes of avoidable emergency department presentations by RACH residents (Hutchinson, Parikh, Tacey, Harvey, & Lim, 2014), and its potential sequalae (e.g., UTI, electrolyte imbalance, increased confusion, falls). In our study, a resident identified with increased aspiration risk based on the CSE, demonstrated functional laryngeal sensation and an effective cough reflex during the mFEES. Thus, unnecessary use of thickened fluids could be avoided.

In another case, mFEES images of food that was silently aspirated (i.e., entered the airway without coughing) were incorporated into the mFEES report. Visual evidence of heightened choking risk with specific foods allowed the resident and his dysphagia care team to make informed swallowing management decisions. This person-specific airway protection assessment may help to reduce the risk of accidental choking, currently reported to be the second leading cause of preventable deaths in adults in RACHs (Ibrahim et al., 2017). Further, a mFEES enabled, person-centered approach that involves individualised and adapted interventions may lead to positive health-related outcomes in adults with multimorbidity and chronic disease (Poitras, Maltais, Bestard-Denommé, Stewart, & Fortin, 2018).

#### Dysphagia Education

Observation of mFEES services and information shared through mFEES reports/recordings offered nursing staff, treating SLPs, residents and families valuable learning opportunities, promoting engagement and informed decision-making by these stakeholders. Healthcare education may increase residents’ perceived agency (Jotterand, Amodio, & Elger, 2016) and reduce anxiety (Spalding, 2003) about dysphagia and its treatment, promoting compliance and satisfaction with care (Fereidouni et al., 2019). Incidental education could also enhance the skills and professional satisfaction of nurses working in RACHs, whose access to professional development may be limited in part due to ubiquitous workforce shortages (Lee et al., 2022). SLPs working in RACHs may also benefit from additional observation, education and training in FEES, an advanced clinical skill, developed through workforce credentialling/training (S. E. Langmore et al., 2022).

#### Collaborative Healthcare

The mFEES service fostered an interdisciplinary approach to dysphagia care, where (i) GPs were required to discuss mFEES referrals with SLPs and approve the procedure, (ii) nursing staff, the referring and the FEES trained SLPs collaborated closely in providing resident care, (iii) and an otolaryngologist was accessible via email/ phone to consult on a needs basis. Interdisciplinary collaboration aligns with calls for innovative care models that facilitate integration between aged care and healthcare services (AS et al., 2020). to reduce fragmentation and support improved healthcare outcomes (AS et al., 2020).

### Acceptability of mFEES Services in RACHs

Many older adults living in RACHs experience cognitive impairment(s), anxiety, pain, and associated behaviours that may challenge their ability to travel to an off-site clinic and to co-operate during an ISA. Our mFEES eliminated possible stressors of off-site travel by providing ISA in the familiar environment of the residents’ bedrooms, incorporating familiar people, objects, and food/drinks. Informal observations echoed findings in the analogous field of mobile radiography that adults with cognitive impairment may feel increasingly safe and calm participating in instrumental assessments that occur in familiar settings (Jensen et al., 2021) (e.g., sitting in their usual chair or bed).

In our study, all adults successfully participated in the mFEES with tailored verbal support and physical compensation. Informal observations suggested that co-operation levels decreased with increasing dementia (cognitive impairment) severity (Jensen et al., 2021).

#### Anxiety Impact on mFEES Participation

Half of the residents in our study reported no anxiety or mild levels of pre-procedural anxiety. Without baseline measures of everyday anxiety, it is difficult to discuss the degree anticipation of the mFEES contributed to anxiety ratings. However, residents with the most severe ratings, had pre-existing heightened anxiety reported by NS. In the absence of mFEES, residents with high baseline anxiety may have found it onerous to travel off-site or to cooperate with an ISA in an unfamiliar environment. We suggest that it is important for healthcare providers to establish a positive connection with residents as part of mFEES to minimise anxiety, ensure resident cooperation, empowerment and well-being, s(Gharibian Adra, Aharonian, & Sibai, 2019). It follows that quality mFEES services in RACHs require allocated time for positive relationship building with residentsand funding structures that reward quality clinical outcomes, in addition to efficient clinical throughput.(Dixit & Sambasivan, 2018).

High level PPE worn by SLP and nurses appeared detrimental to effective communication with adults with pre-existing hearing impairment and/or dementia, potentially contributing to resident anxiety. However, PPE protocols were prioritised to minimise the risks of COVID-19 exposure.

#### Procedural Comfort

Most residents reported low levels of discomfort during the mFEES. Our findings align with previous studies where the average pain level reported by participants ranged from (i) 2.00-3.46/ 10 for endoscope insertion and (ii) 2.52-3.00/ 10 for the actual FEES procedure (Farneti, Fattori, & Bastiani, 2017; Fife et al., 2015). Interestingly, residents who reported higher levels of pre-procedural anxiety also reported higher pain levels. In our study, lubrication without anesthetic was used during endoscope insertion to avoid (i) disrupting peripheral sensation and contingent motor aspects of the swallow, (ii) the low risk of anesthetic complications (Kamarunas, McCullough, Guidry, Mennemeier, & Schluterman, 2014). However, some studies suggest that the use of anesthetic can improve comfort ratings in older adults (> 60 years), without significantly impacting swallow function (Fife et al., 2015; O’Dea et al., 2015). It may be helpful to consider anesthetic application on an individual basis as part of future mFEES services, particularly in older adults with: (i) pre-existing heightened anxiety (due to the recognised reciprocal relationship between pain and anxiety in upper endoscopy)(Lauriola et al., 2019); or known nasal/pharyngeal sensitivity.

### Limitations

Participant recruitment was challenged by COVID-19 related intermittent RACHs lockdowns. SLPs working in RACHs reported stricter referral triaging practices and a decrease in referral numbers, further reducing the pool of potential study participants.

While residents in our study represented a range of age groups, medical aetiologies, genders and geographical localities, the generalizability of our results may be un-representative due to incidental participant recruitment and the relatively small sample size. For example, the post-mFEES survey was completed by only four medical decision-makers because some residents did not require MDMs, while others had government appointed MDMs or MDMs who were unavailable to attend the mFEES.

Due to malfunction of the Telepack X ED motherboard, one mFEES video file could not be downloaded to a flash drive for analysis. The mFEES report was written based on real-time observations by the R-SLP. For all subsequent procedures, two endoscopy units were available at each resident’s bedside in case of equipment malfunction.

Further research with larger participant numbers is needed to explore the impact of mFEES on residents, their families, healthcare providers and community healthcare resources. For example, residents’ opinions, swallowing related outcomes (e.g., clinical risk measures including pneumonia, choking and emergency department admission rates; QOL; nutrition; and hydration levels etc.), fiscal outcomes (e.g., formal cost analysis/comparison of mFEES vs off-site ISA service models), and mFEES complication rates would be useful in designing financially viable, high quality future mFEES services.

## Conclusion

Improved healthcare for older adults living in RACHs is an international priority (Barker et al., 2018). Adults with OD do not have easy access to timely ISA, a standard component of swallowing care available to adults in other healthcare settings. In this feasibility study we trialled a mFEES service model providing onsite, person-centered ISA to adults in RACHs in Australia. Our findings suggest that mFEES is a feasible and well tolerated service with potential to reduce clinical risks of OD, and to enhance the quality of swallowing care available to older adults in RACHs. Information obtained during the mFEES can be used by healthcare staff, residents, and their families to make increasingly timely, informed, and tailored decisions about swallowing management. Further, our mFEES service supported an integrated healthcare model and offered learning opportunities for RACHs staff (including SLP, nursing, carers), family members, and residents. Our study indicates potential for mFEES in RACHs and the need for further research with larger participant numbers to further explore the effect of mFEES on clinical risk, healthcare outcomes, and the cost of high-quality swallowing care.

## Data Availability

The data that support the findings of this study are available on request from the corresponding author. The data are not publicly available due to privacy or ethical restrictions.

## Declaration of Conflicting Interests

The Authors declare that there is no conflict of interest

## Funding

This scoping review was conducted as part of a PhD research program. O.B. is receiving Australian Government Research Training Scholarship funding through the University of Melbourne to participate in this research program.

A.V. holds a fellowship from the National Health and Medical Research Council, Australia (no. 1135683).

Sue Cotton is supported by a National Health and Medical Council Senior Research Fellowship (SRF, no. APP1136344).

## Acknowledgments

We would like to thank and acknowledge residential aged care and speech pathology service providers who participated in this research project. The names of specific facilities and healthcare staff have been withheld to protect participant confidentiality.

We are grateful to Patricio Rojas (Clinical Support & EndoProtect 1 Specialist from KARL STORZ Endoscopy Australia Pty Ltd), Jennifer Rennie and Alicja Britten (Product Consultants, Visualization Ambu Australia Pty. Ltd) for their technical support, problem solving and expertise.

## Conflict of Interest

The authors declare no conflicts of interest.

## Ethical Approval

Ethical approval was granted by The University of Melbourne Human Research and Ethics Committee (reference number 021-13387-13943-2, 08/02/21).

## Patient consent statement

All participants in this study, or if appropriate, their legally appointed medical decision makers, read the study Plain Language Statement and signed a Consent Form before inclusion in the study.

## Notes

### Competing Interest Statement

The authors have declared no competing interest.

### Clinical Trial

Australian New Zealand Clinical Trials Registry (ANZCTR): ACTRN12622001369718

## References

Alfred Health. (2021, Oct 15). Specialist clinic & outpatient information. https://www.alfredhealth.org.au/covid-19/specialist-clinic-and-outpatient-information#:~:text=As%20part%20of%20Alfred%20Health’s,more%20limited%20at%20this%20time

Australian Nursing and Midwidery Federation. (2020). Submission of the Australian Nursing and Midwifery Federation in Relation to the Impact of COVID-19 in Aged Care. https://www.anmf.org.au/resources/submissions

Baijens, L. W. J., Clavé, P., Cras, P., Ekberg, O., Forster, A., Kolb, G. F., Leners, J. C., Masiero, S., Mateos-Nozal, J., Ortega, O., Smithard, D. G., & Walshe, M. (2016). European society for swallowing disorders - European union geriatric medicine society white paper: Oropharyngeal dysphagia as a geriatric syndrome. Clinical Interventions in Aging, 11, 1403–1428. http://doi.org/10.2147/CIA.S107750

Barczi, S. R., Sullivan, P. A., & Robbins, J. A. (2000). How Should Dysphagia Care of Older Adults Differ? Establishing Optimal Practice Patterns. Semin Speech Lang, 21(04), 0347–0364. http://doi.org/10.1055/s-2000-8387

Barker, R. O., Craig, D., Spiers, G., Kunonga, P., & Hanratty, B. (2018). Who Should Deliver Primary Care in Long-term Care Facilities to Optimize Resident Outcomes? A Systematic Review. Journal of the American Medical Directors Association, 19(12), 1069–1079. https://doi.org/10.1016/j.jamda.2018.07.006

Birchall, O., Bennett, M., Lawson, N., Cotton, S. M., & Vogel, A. P. (2021a). Instrumental Swallowing Assessment in Adults in Residential Aged Care Homes: A Scoping Review. J Am Med Dir Assoc, 22(2), 372–379.e376. https://doi.org/10.1016/j.jamda.2020.08.028

Birchall, O., Bennett, M., Lawson, N., Cotton, S. M., & Vogel, A. P. (2021b). The Role of Instrumental Swallowing Assessment in Adults in Residential Aged Care Homes: A National Modified Delphi Survey Examining Beliefs and Practices. Dysphagia, 37(3), 510–522. https://doi.org/10.1007/s00455-021-10296-2

Birchall, O., Bennett, M., Lawson, N., Cotton, S. M., & Vogel, A. P. (2022). Instrumental swallowing assessment in adults in residential aged care homes: practice patterns and opportunities. Australas J Ageing, 00:1–10. https://doi.org/10.1111/ajag.13122

Cao, X., Yumul, R., Elvir Lazo, O. L., Friedman, J., Durra, O., Zhang, X., & White, P. F. (2017). A novel visual facial anxiety scale for assessing preoperative anxiety. Plos One, 12(2), e0171233. https://doi.org/10.1371/journal.pone.0171233

Chalmers, J., & Johnson, V. (2004). Evidence-based protocol: oral hygiene care for functionally dependent and cognitively impaired older adults. Journal Of Gerontological Nursing, 30(11), 5–12. https://doi.org/10.3928/0098-9134-20041101-06

Chen, S., Kent, B., & Cui, Y. (2021). Interventions to prevent aspiration in older adults with dysphagia living in nursing homes: a scoping review. BMC Geriatrics, 21(1), 429. https://doi.org/10.1186/s12877-021-02366-9

Cimoli, M., & Sweeney, J. (2012). Fibreoptic Endoscopic Evaluation of Swallowing (FEES): Models of service delivery and approaches to training. J Clin Pract Speech Lang Pathol, 14(1), 18–24.

Crary, M. A., Mann, G. D. C., & Groher, M. E. (2005). Initial psychometric assessment of a functional oral intake scale for dysphagia in stroke patients. Archives of Physical Medicine & Rehabilitation, 86(8), 1516–1520. https://doi.org/10.1016/j.apmr.2004.11.049

Dixit, S. K., & Sambasivan, M. (2018). A review of the Australian healthcare system: A policy perspective. SAGE Open Med, 6, 2050312118769211. https://doi.org/10.1177/2050312118769211

Donzelli, J., Brady, S., Wesling, M., & Craney, M. (2003). Predictive value of accumulated oropharyngeal secretions for aspiration during video nasal endoscopic evaluation of the swallow. Annals of Otology, Rhinology & Laryngology, 112(5), 469–475. https://doi.org/10.1177/000348940311200515

Eagar K, W. A., Snoek M, Kobel C, Loggie C and Gordon R, & Centre for Health Service Development, A. H. S. R. I., University of Wollongong. (2019). How Australian residential aged care staffing levels compare with international and national benchmarks. https://agedcare.royalcommission.gov.au/sites/default/files/2019-12/research-paper-1.pdf

Farneti, D., Fattori, B., & Bastiani, L. (2017). The endoscopic evaluation of the oral phase of swallowing (Oral-FEES, O-FEES): a pilot study of the clinical use of a new procedure. Acta otorhinolaryngologica Italica : organo ufficiale della Societa italiana di otorinolaringologia e chirurgia cervico-facciale, 37(3), 201–206. https://doi.org/10.14639/0392-100X-1126

Fereidouni, Z., Sabet Sarvestani, R., Hariri, G., Kuhpaye, S. A., Amirkhani, M., & Kalyani, M. N. (2019). Moving Into Action: The Master Key to Patient Education. J Nurs Res, 27(1), 1–8. https://doi.org/10.1097/jnr.0000000000000280

Fife, T., Butler, S., Langmore, S., Lester, S., Wright, S. C., Kemp, S., Grace-Martin, K., & Lintzenich, C. (2015). Use of topical nasal anesthesia during flexible endoscopic evaluation of swallowing in dysphagic patients. Annals of Otology, Rhinology, and Laryngology, 124(3), 206–211. https://doi.org/10.1177/0003489414550153

Gephart, S. M., Effken, J. A., McGrath, J. M., & Reed, P. G. (2013). Expert consensus building using e-Delphi for necrotizing enterocolitis risk assessment. J Obstet Gunecol Neonatal Nurs, 42(3), 332–347. https://doi.org/10.1111/1552-6909.12032

Gharibian Adra, M., Aharonian, Z., & Sibai, A. M. (2019). Exploring resident-staff relationships in nursing homes in Lebanon. Int J Qual Stud Health Well-being, 14(1), 1688605–1688605. https://doi.org/10.1080/17482631.2019.1688605

Gilbert, A. S., E, Owusu-Addo, E., Feldman, P., Mackell, P., Garratt, S. M., & Brijnath, B. (2020, August 13). Models of Integrated Care, Health and Housing: Report prepared for the Royal Commission into Aged Care Quality and Safety, National Ageing Research Institute. https://agedcare.royalcommission.gov.au/sites/default/files/2020-08/Research%20Paper%207%20-%20Models%20of%20integrated%20care%2C%20health%20and%20housing.pdf

Grabowski, D. C., & Mor, V. (2020). Nursing Home Care in Crisis in the Wake of COVID-19. JAMA, 324(1), 23–24. https://doi.org/10.1001/jama.2020.8524

Harris, P. A., Taylor, R., Minor, B. L., Elliott, V., Fernandez, M., O’Neal, L., McLeod, L., Delacqua, G., Delacqua, F., Kirby, J., & Duda, S. N. (2019). The REDCap consortium: Building an international community of software platform partners. Journal of Biomedical Informatics, 95, 103208. https://doi.org/10.1016/j.jbi.2019.103208

Harris, P. A., Taylor, R., Thielke, R., Payne, J., Gonzalez, N., & Conde, J. G. (2009). Research electronic data capture (REDCap)—A metadata-driven methodology and workflow process for providing translational research informatics support. Journal of Biomedical Informatics, 42(2), 377–381. https://doi.org/10.1016/j.jbi.2008.08.010

Hase, T., Miura, Y., Nakagami, G., Okamoto, S., Sanada, H., & Sugama, J. (2019). Food bolus-forming ability predicts incidence of aspiration pneumonia in nursing home older adults: A prospective observational study. Journal of Oral Rehabilitation, 47(1), 53–60. https://doi.org/10.1111/joor.12861

Hutchinson, A. F., Parikh, S., Tacey, M., Harvey, P. A., & Lim, W. K. (2014). A longitudinal cohort study evaluating the impact of a geriatrician-led residential care outreach service on acute healthcare utilisation. Age and Ageing, 44(3), 365–370. https://doi.org/10.1093/ageing/afu196

Ibrahim, J. E., Bugeja, L., Willoughby, M., Bevan, M., Kipsaina, C., Young, C., Pham, T, & Ranson, D. L. (2017). Premature deaths of nursing home residents: an epidemiological analysis. Medical Journal of Australia, 206(10), 442–447. https://doi.org/10.5694/mja16.00873

Jensen, J. M., Andersen, P. A. B., Kirkegaard, L., Larsen, N., Most, W., Nielsen, D., & Precht, H. (2021). Exploring the patient perspectives of mobile X-ray in nursing homes – A qualitative explorative pilot study. Radiography, 27(2), 279–283. https://doi.org/10.1016/j.radi.2020.08.009

Jotterand, F., Amodio, A., & Elger, B. S. (2016). Patient education as empowerment and self-rebiasing. Medicine, Health Care and Philosophy, 19(4), 553–561. https://doi.org/10.1007/s11019-016-9702-9

Jukic Peladic, N., Orlandoni, P., Dell’Aquila, G., Carrieri, B., Eusebi, P., Landi, F., Volpato, S., Zuliani, G., Lattanzio, F., & Cherubini, A. (2018). Dysphagia in Nursing Home Residents: Management and Outcomes. J Am Med Dir Assoc, 20(2), 147–151. https://doi.org/10.1016/j.jamda.2018.07.023

Kamarunas, E. E., McCullough, G. H., Guidry, T. J., Mennemeier, M., & Schluterman, K. (2014). Effects of topical nasal anesthetic on fiberoptic endoscopic examination of swallowing with sensory testing (FEESST). Dysphagia, 29(1), 33–43. https://doi.org/10.1007/s00455-013-9473-x

Kelly, A. M., McLaughlin, C., Wallace, S., Hales, P., Stewart, C., Leathley, C., & Cunningham, R. (2015). Fibreoptic Endoscopic Evaluation of Swallowing (FEES): The role of speech and language therapy. Royal College of Speech and Language Therapists Position Paper. https://docplayer.net/20821980-Position-paper-fibreoptic-endoscopic-evaluation-of-swallowing-fees-the-role-of-speech-and-language-therapy.html

Langmore, S., & Aviv, J. E. (2001). Endoscopic Procedures to Evaluate Oropharyngeal Swallowing. In S. Langmore (Ed.), Endoscopic Evaluation and Treatment of Swallowing Disorders (pp. 73–100). New York: Thieme.

Langmore, S. E., Scarborough, D. R., Kelchner, L. N., Swigert, N. B., Murray, J., Reece, S., Cavanagh, T., Harrigan, L. C., Scheel, R., Gosa, M. M., & Rule, D. K. (2022). Tutorial on Clinical Practice for Use of the Fiberoptic Endoscopic Evaluation of Swallowing Procedure With Adult Populations: Part 1. Am J Speech Lang Pathol, 31(1), 163–187. https://doi.org/10.1044/2021_ajslp-20-00348

Lauriola, M., Tomai, M., Palma, R., La Spina, G., Foglia, A., Panetta, C.,… Pontone, S. (2019). Intolerance of Uncertainty and Anxiety-Related Dispositions Predict Pain During Upper Endoscopy. Frontiers in psychology, 10, 1112–1112. doi:10.3389/fpsyg.2019.01112

Leder, S. B., & Neubauer, P. D. (2016). The Yale Pharyngeal Residue Severity Rating Scale: Springer. https://link.springer.com/book/10.1007/978-3-319-29899-3

Lee, H. Y., Short, S., Lee, M.-J., Jeon, Y.-H., Park, E., & Chin, Y.-R. (2022). Improving the quality of long-term care services in workforce dimension: expert views from Australia and South Korea. Archives of Public Health, 80(1), 112. https://doi.org/10.1186/s13690-022-00872-9

Mehrotra, A., Chernew, M. E., Linetsky, D., Hatch, H., & Cutler, D. A. (2020, May 19). The Impact of the COVID-19 Pandemic on Outpatient Visits: A Rebound Emerges. https://www.commonwealthfund.org/publications/2020/apr/impact-covid-19-outpatient-visits

Muschol, J., & Gissel, C. (2021). COVID-19 pandemic and waiting times in outpatient specialist care in Germany: an empirical analysis. BMC Health Services Research, 21(1), 1076. https://doi.org/10.1186/s12913-021-07094-9

Nogueira, D., & Reis, E. (2013). Swallowing disorders in nursing home residents: how can the problem be explained? Clinical Interventions in Aging, 8, 221–227. https://doi.org/10.2147/CIA.S39452

O’Dea, M., Langmore, S., Krisciunas, G., Walsh, M., Zanchetti, L., Scheel, R., McNally, E., Kaneoka, A. S., Guarino, A. J., & G Butler, S. (2015). Effect of Lidocaine on Swallowing During FEES in Patients With Dysphagia 124(7). https://doi.org/10.1177/0003489415570935

Poitras, M.-E., Maltais, M.-E., Bestard-Denommé, L., Stewart, M., & Fortin, M. (2018). What are the effective elements in patient-centered and multimorbidity care? A scoping review. BMC Health Services Research, 18(1), 446. https://doi.org/10.1186/s12913-018-3213-8

Prvu Bettger, J., Thoumi, A., Marquevich, V., De Groote, W., Rizzo Battistella, L., Imamura, M., Ramos, V. D., Wang, N., Dreinhoefer, K. E., Mangar, A., Ghandi, d. b. c., Ng, Y. S., Lee, K. H., Ming, J. T. W., Pua, Y. H., Inzitari, M., Mmbaga, B. T., Shayo, M. J., Brown, D. A., Carvalho, M., Oh-Park, M., & Stein, J. (2020). COVID-19: maintaining essential rehabilitation services across the care continuum. BMJ Glob Health, 5(5). https://doi.org/10.1136/bmjgh-2020-002670

Rogus Pulia, N., Wirth, R., & Sloane, P. D. (2018). Dysphagia in Frail Older Persons: Making the Most of Current Knowledge. J Am Med Dir Assoc, 19(9), 736–740. https://doi.org/10.1016/j.jamda.2018.07.018

Rosenbek, J. C., Robbins, J., Roecker, E. B., Coyle, J. L., & Wood, J. L. (1996). A penetration-aspiration scale. Dysphagia, 11, 93–98. http://dx.doi.org/10.1007/BF00417897

Rosenvinge, S. K., & Starke, I. D. (2005). Improving care for patients with dysphagia. Age and Ageing, 34(6), 587–593. https://doi.org/10.1093/ageing/afi187

Spalding, N. J. (2003). Reducing anxiety by pre-operative education: make the future familiar. Occupational Therapy International, 10(4), 278–293. https://doi.org/10.1002/oti.191

Speech Pathology Australia. (2019) Clinical guideline: Flexible Endoscopic Evaluation of Swallowing (FEES) 2019. https://www.speechpathologyaustralia.org.au/SPAweb/Members/Practice_Guidelines/SPAweb/Members/Clinical_Guidelines/Clinical_Guidelines.aspx?hkey=0fc81470-2d6c-4b17-90c0-ced8b0ff2a5d. Accessed March 3, 2022.

Steele, C. M., Greenwood, C., Ens, I., Robertson, C., & Seidman Carlson, R. (1997). Mealtime difficulties in a home for the aged: not just dysphagia. Dysphagia, 12(1), 43–50. https://doi.org/10.1007/pl00009517

Takahashi, N., Kikutani, T., Tamura, F., Groher, M., & Kuboki, T. (2012). Videoendoscopic assessment of swallowing function to predict the future incidence of pneumonia of the elderly. Journal of Oral Rehabilitation, 39(6), 429–437. https://doi.org/10.1111/j.1365-2842.2011.02286.x

van der Maarel-Wierink, C.D., Vanobbergen, J.N., Bronkhorst, E.M., Schols, J.M., de Baat, C. (2011) Risk factors for aspiration pneumonia in frail older people: a systematic literature review. J Am Med Dir Assoc. 2011 Jun;12(5), 344–54. https://doi: 10.1016/j.jamda.2010.12.099. Epub 2011 Mar 21. PMID: 21450240

Wakabayashi, H. (2014). Presbyphagia and Sarcopenic Dysphagia: Association between Aging, Sarcopenia, and Deglutition Disorders. The Journal of frailty & aging, 3 2, 97–103. https://doi.org/10.14283/jfa.2014.8

Xu, H., Intrator, O., & Bowblis, J. R. (2020). Shortages of Staff in Nursing Homes During the COVID-19 Pandemic: What are the Driving Factors? J Am Med Dir Assoc, 21(10), 1371–1377. https://doi.org/10.1016/j.jamda.2020.08.002

