## Appendix 1 - 8 for "Mobile instrumental assessment of swallowing in residential aged care homes"

**Appendix 1.** Materials provided by the Residential Aged Care Homes for the mFEES procedure.

- plastic examination gloves

- 4 x teaspoons

- 1 x tablespoon

- 1 x fork

- 4 x glasses of cordial (200m each)

- 1 x plate

- 1 cup of tea/ coffee (on request)

- 1 dry cracker or biscuit

- 1 slice of bread with butter

- 1 banana

- canned diced fruit (eg. peach or apricot)

- 1 tub of yogurt

- food/ fluids in addition to materials specified above in the following circumstances: it is medically indicated due to the resident’s intolerance(s)/ allergy(ies) or medical condition(s); the examiner (researcher) believes that it is appropriate to trial foods/fluids that are familiar and/or problematic for the participant; the participant expresses a preference for specific food/ fluids

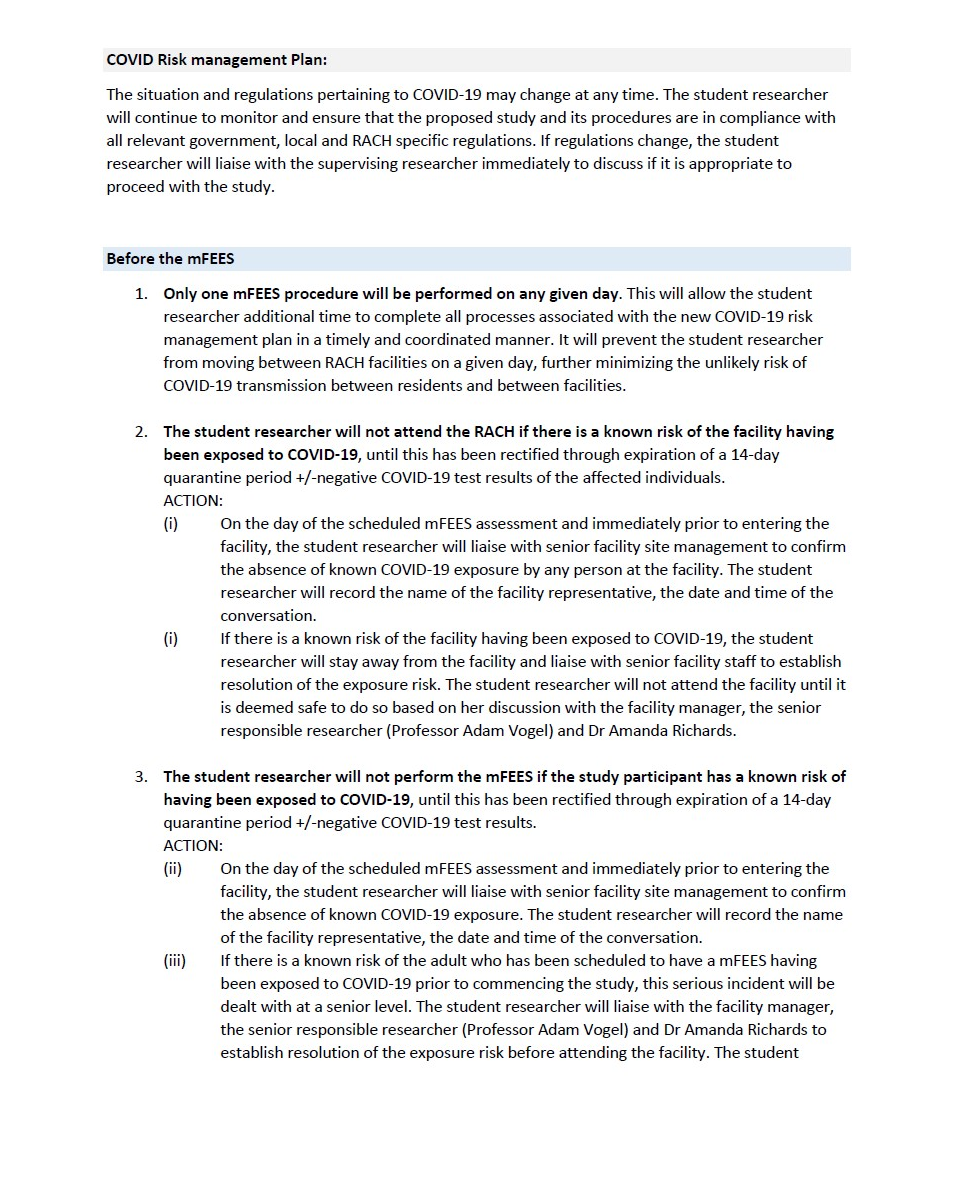
**Appendix 2.** COVID-19 mFEES safety protocol

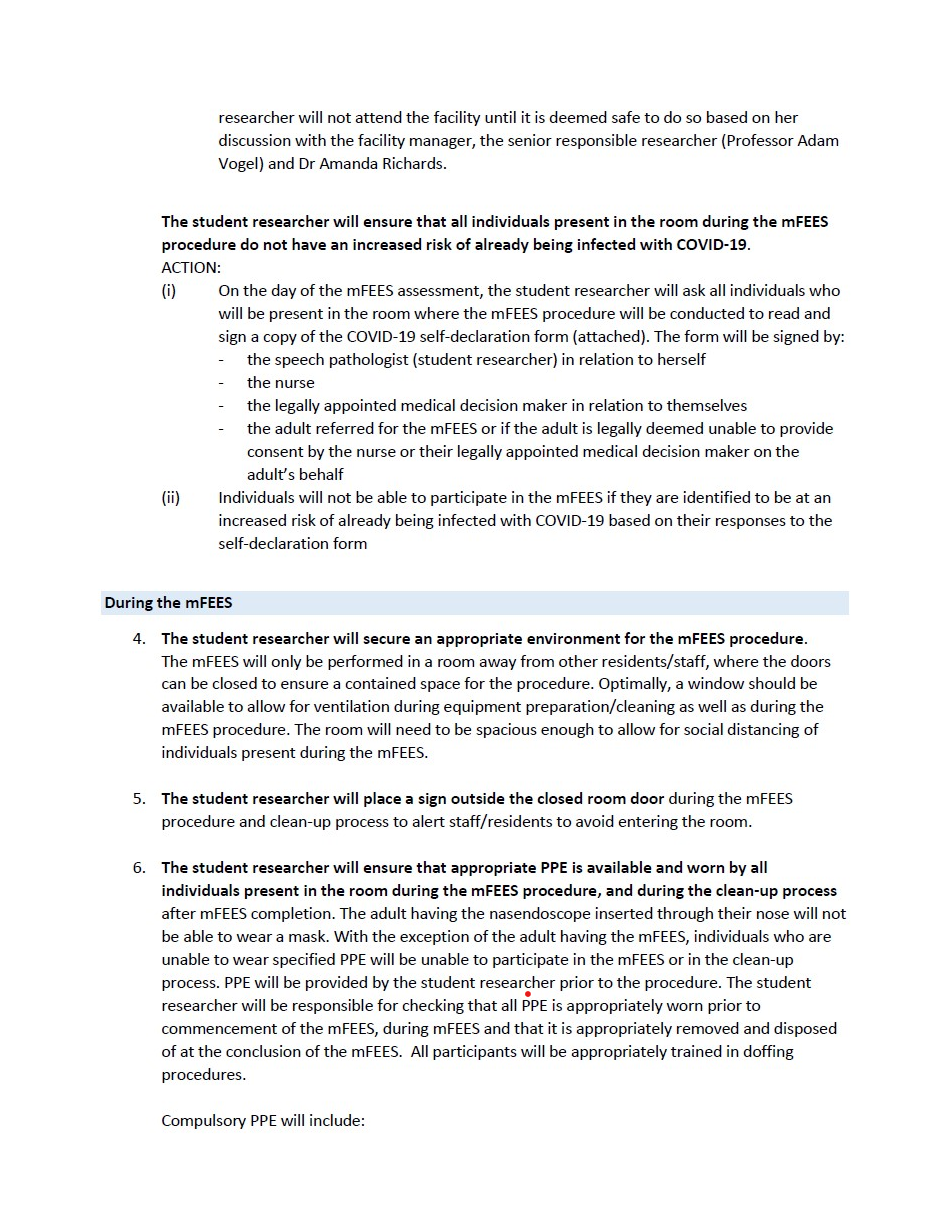

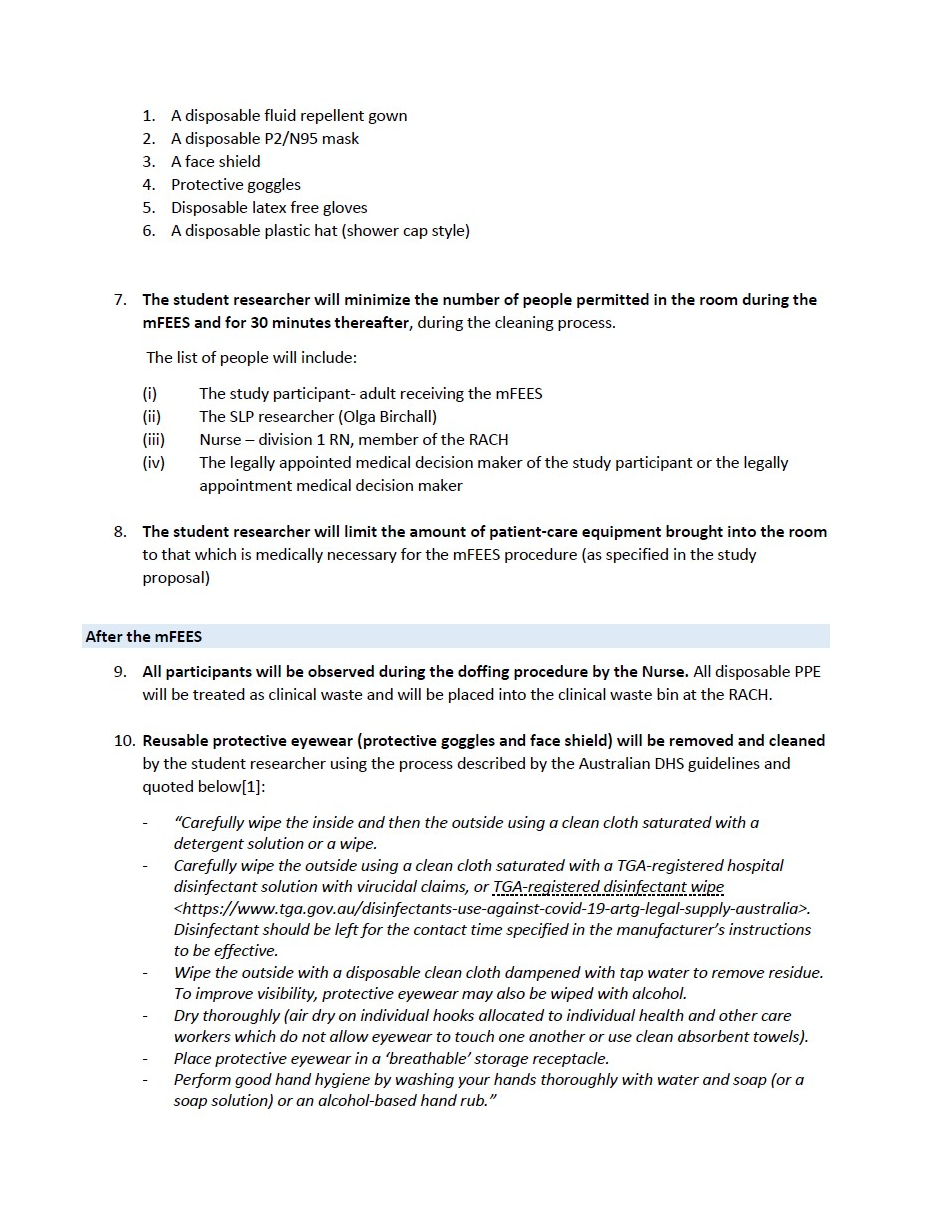

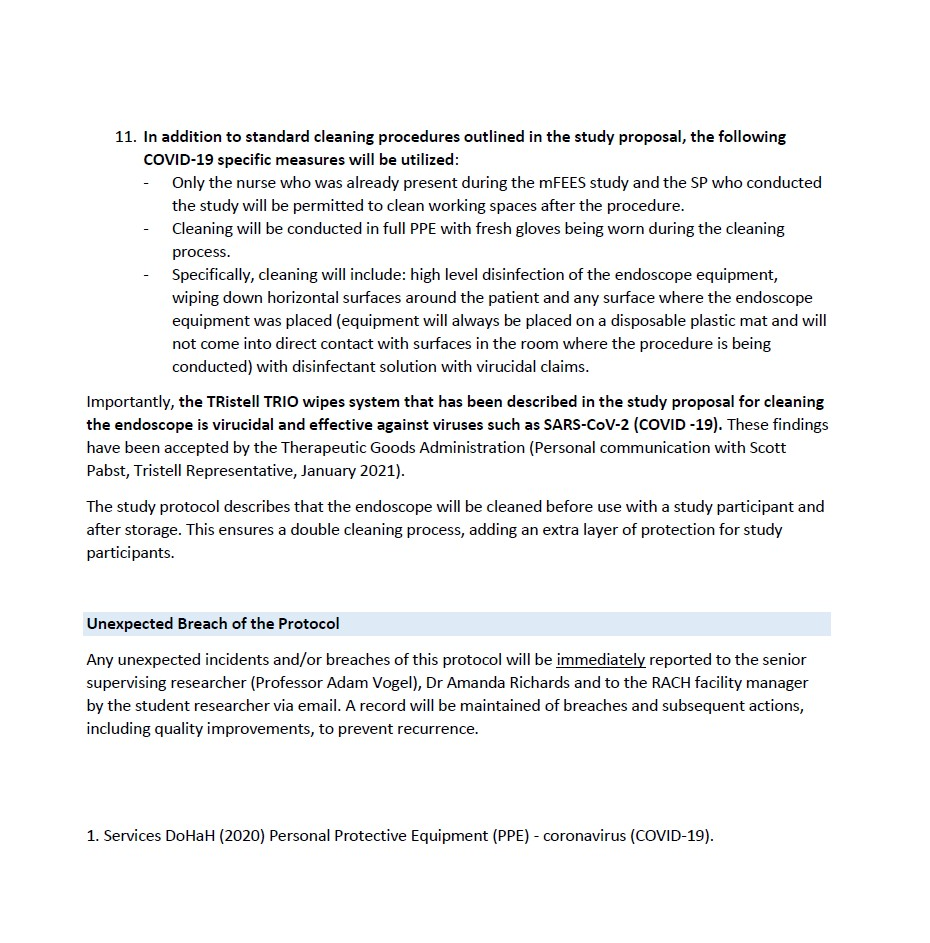

1. Department of Health and Human Services. Personal Protective Equipment (PPE) - coronavirus (COVID-19) [Press release]. Updated 2020. Accessed Feb 02, (2021). Available from: <https://www.dhhs.vic.gov.au/personal-protective-equipment-ppe-covid-19>

**Appendix 3**. mFEES resident inclusion/exclusion criteria

| **Inclusion Criteria** | **Exclusion Criteria** |
| --- | --- |
| - 18 years or older - Resident identified with potential to benefit from FEES by their treating speech pathologist | - unstable cardiac condition - unstable neurological condition e.g., in the process of a suspected stroke - moderate-to-severe behavioural agitation, distress or aggression - oxygen requirements exceed levels provided through nasal prongs ( > 5L/ min) - severe movement disorder with hyperkinetic movements - history or risk of severe epistaxis - history of recurrent vasovagal episodes - recent facial fractures or surgery - known severe hypersensitivity of the nose and/ or nasopharynx and/ or oropharynx - active infectious disease/ condition requiring physical isolation from other residents - bilateral nasal obstruction - base of skull fracture - identified by the treating speech pathologist as not having an adequate understanding of English, due to non-English speaking background, to consider the plain language statement and to make an informed decision about study participation - medically ‘highly vulnerable’ as indicated by the treating GP/ Geriatrician in discussion with the research speech pathologist* |

*In addition to the above criteria, the student researcher, could use her clinical judgement to exclude participants who she suspected were vulnerable and inappropriate for assessment due to multiple medical comorbidities. This decision would first be discussed with the treating GP or Geriatrician.

**Appendix 4.** Equipment used to provide mFEES in RACHs

| **FEES equipment** | **Reusable set-up** | **Disposable set-up** |
| --- | --- | --- |
|  | Karl Storz Monitor (TP 100 – Tele Pack X LED) | Ambu® a View^TM^ 2 Advance portable monitor (touch screen) |
|  | Strobo-Video-Rhino-Laryngoscope  Light source/cable | Ambu® a Scope^TM^ 4 Rhino Laryngo Slim (disposable nasendoscope) |
|  | Power cord | Charging cable (optional) |
|  | Plug-in microphone | External sound recording equipment |
|  | Silicone keyboard (USB connection) |  |
|  | External hard drive (Toshiba 32 GB USB 2.0 Flash Drive) |  |
|  | Leak testing equipment (manometer) |  |
| **Procedural Consumables** | KY lubricant jelly (for nasendoscope lubrication) | |
|  | Alcohol square wipes (for nasendoscope lens cleaning) | |
|  | Disposable waterproof hospital mats (upper soft cotton layer and lower waterproof plastic layer) for ‘clean’ and ‘dirty’ endoscope zones | |
|  | Food dye (blue, green, and white)  Plastic apron (optional for residents to prevent clothes staining) | |
| **PPE** | Plastic gloves | |
|  | Plastic hair covering | |
|  | Waterproof hospital gowns | |
|  | Plastic goggles | |
|  | Face shields | |
|  | N95 masks | |
| **Cleaning products** | Tristel Trio Wipes System (only for reusable rhinolaryngoscope cleaning) | |
|  | Surface disinfectant wipes (for wiping flat surfaces) | |

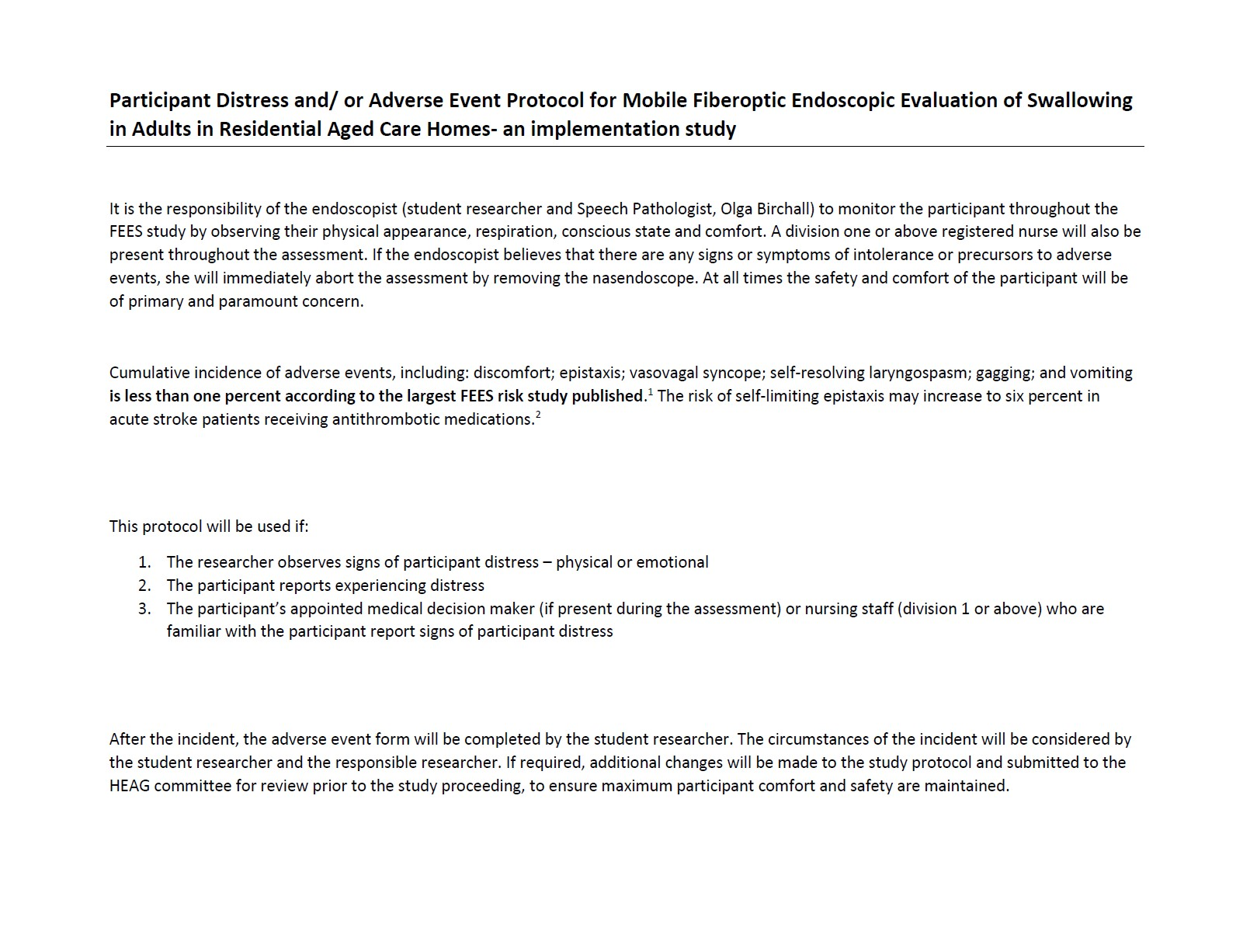
**Appendix 5.** mFEES adverse event protocol

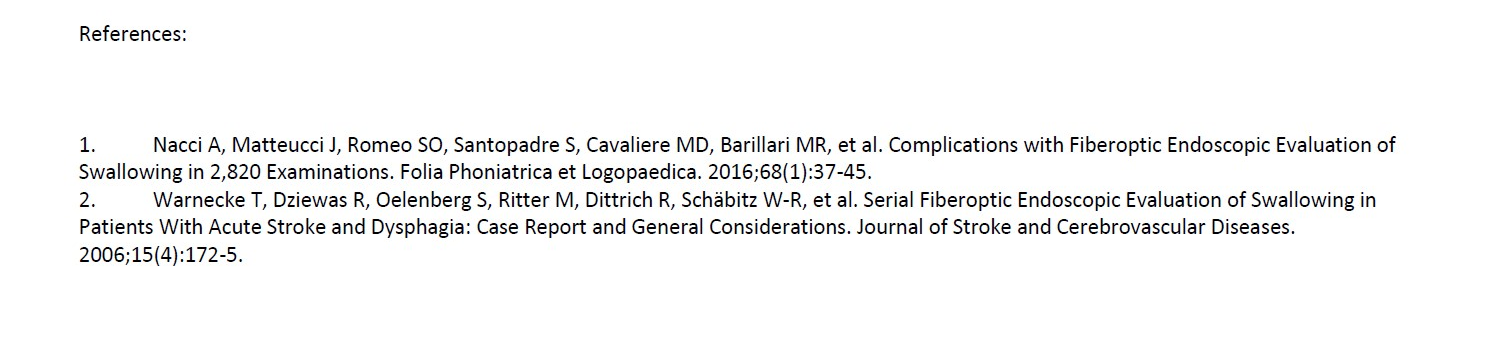

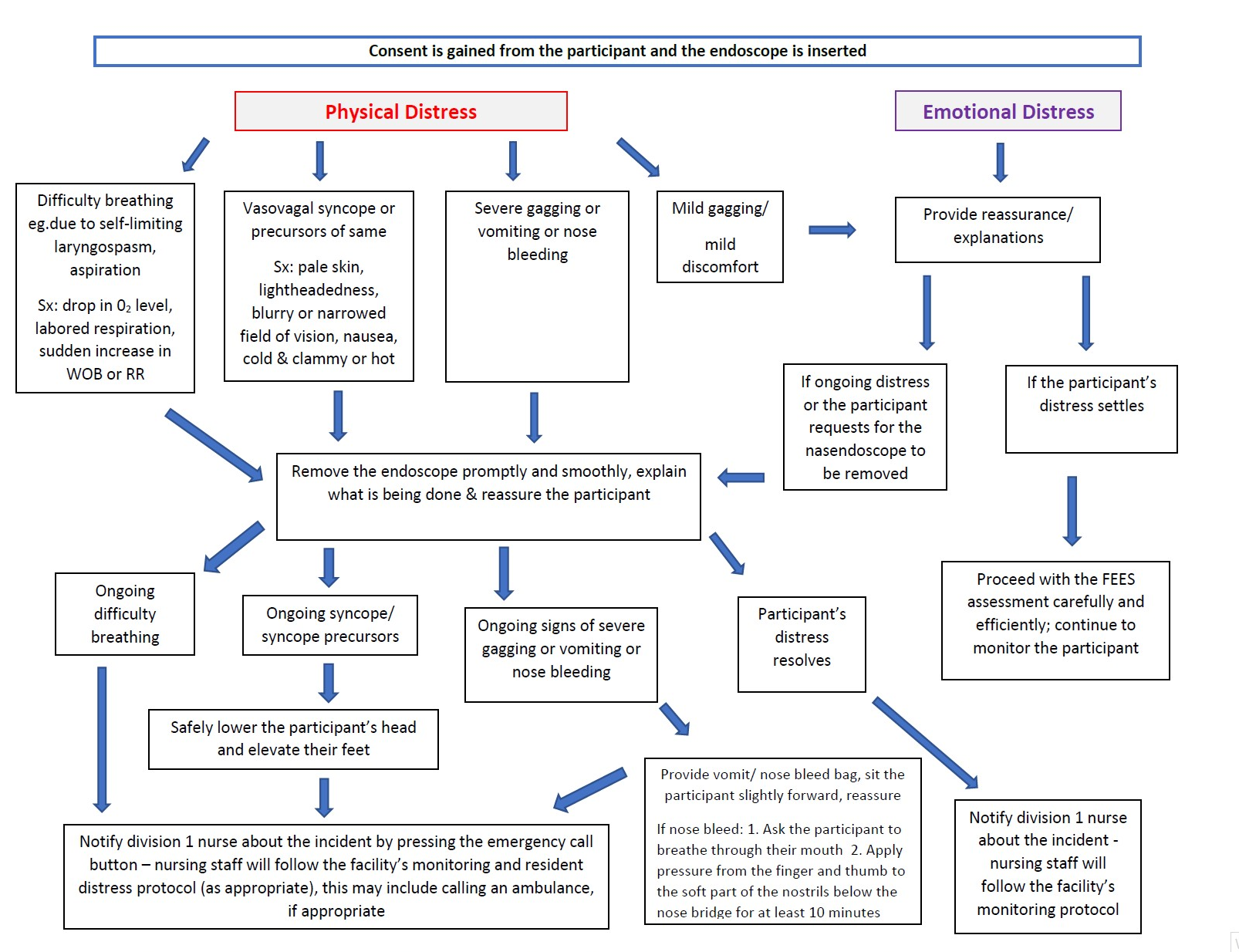

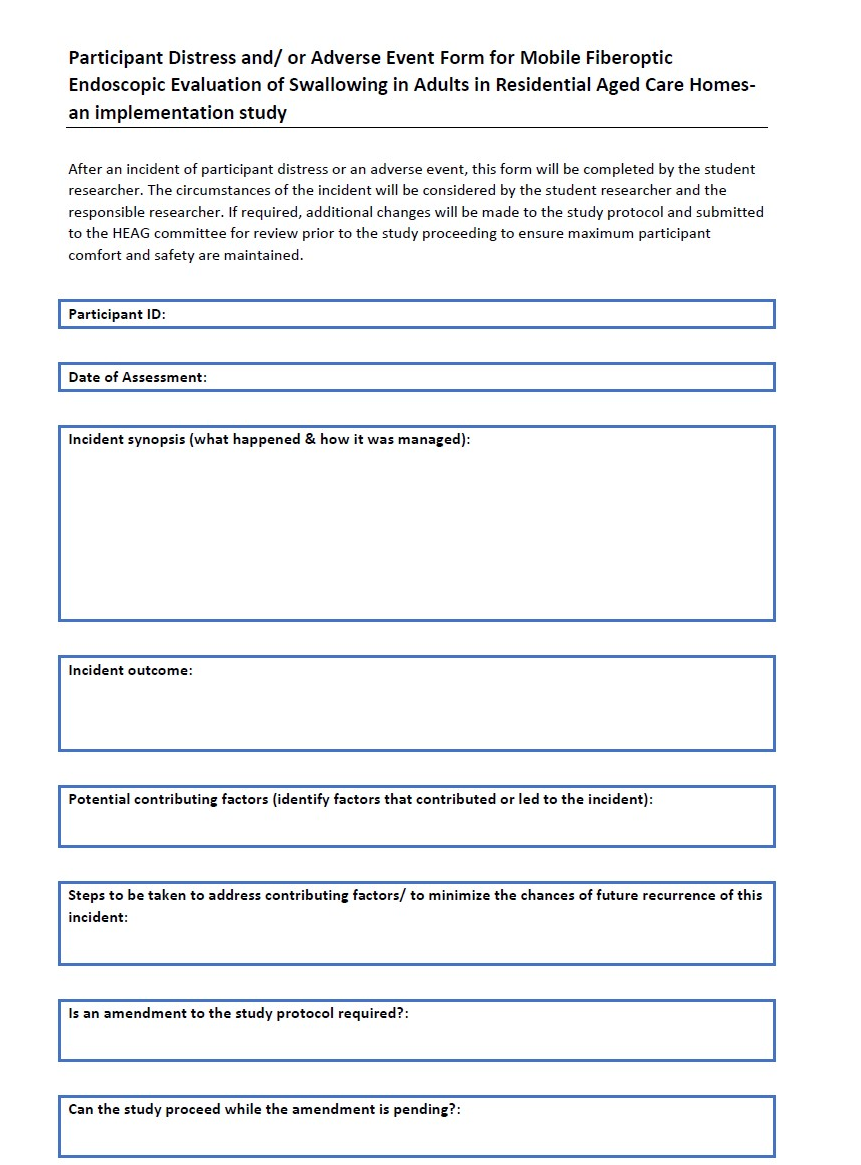

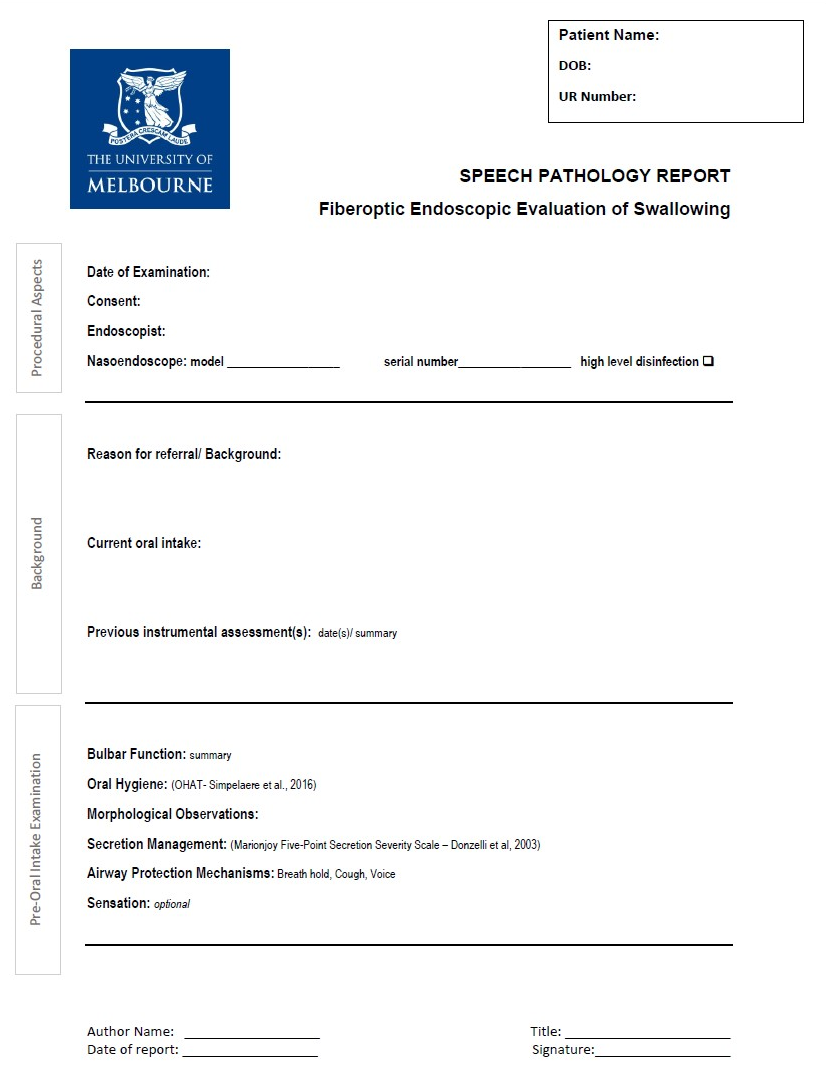
**Appendix 6**. mFEES report proforma

*Removed institution* *identification for confidentiality*

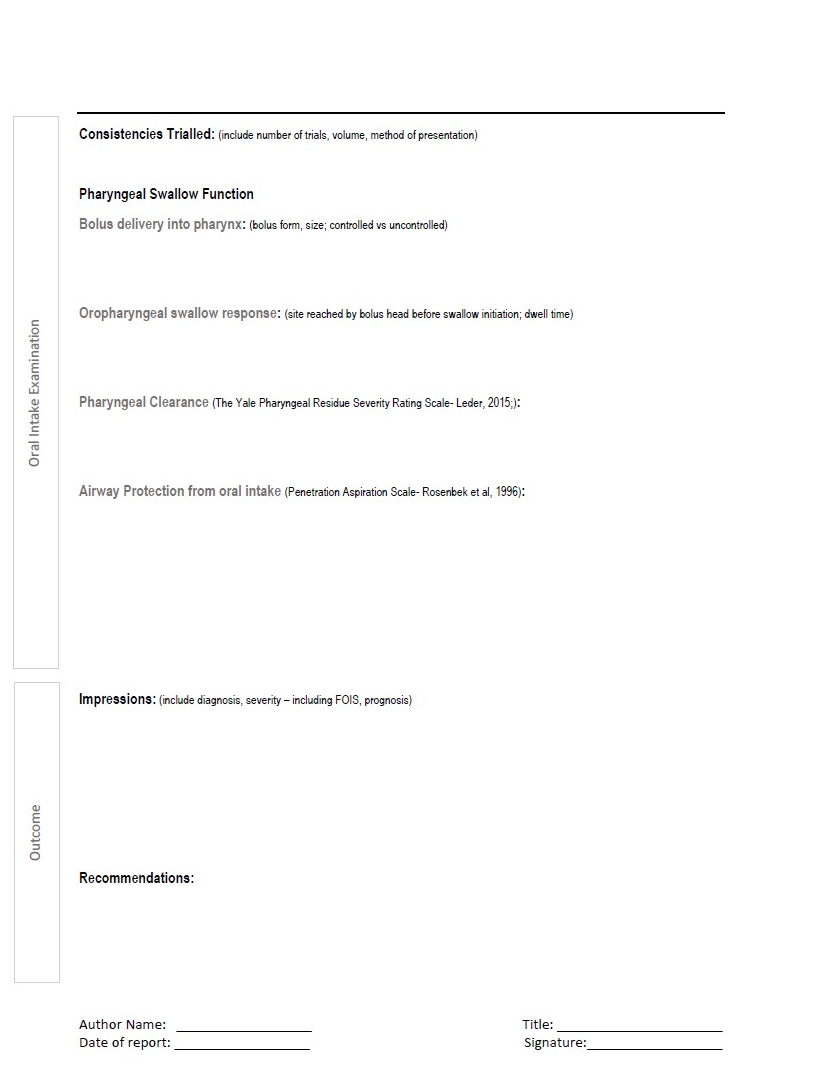

Based on FEES report proforma developed at the University of Melbourne for the Swallowing Outpatient Clinic 2019.

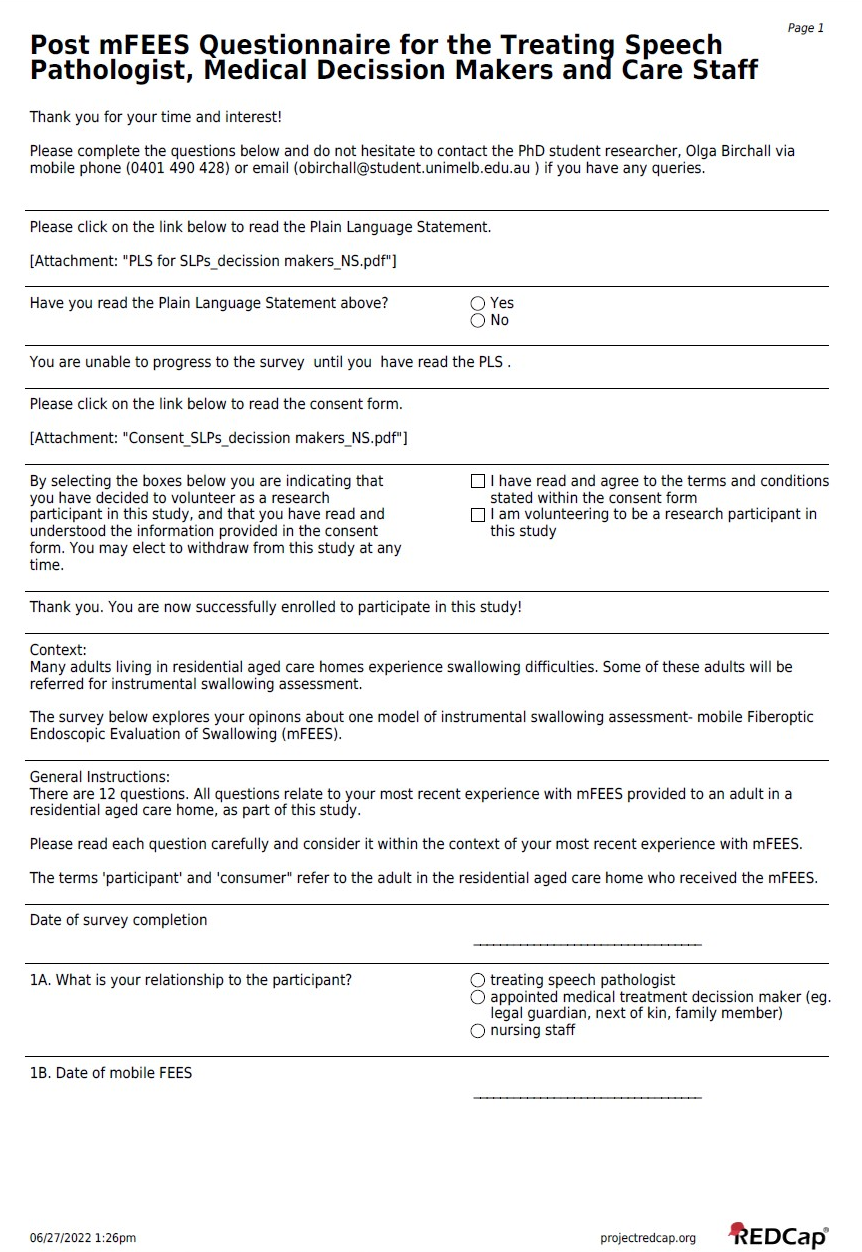
**Appendix 7**. Post-mFEES survey for referring SLPs, nursing staff and legally appointed medical decision makers.

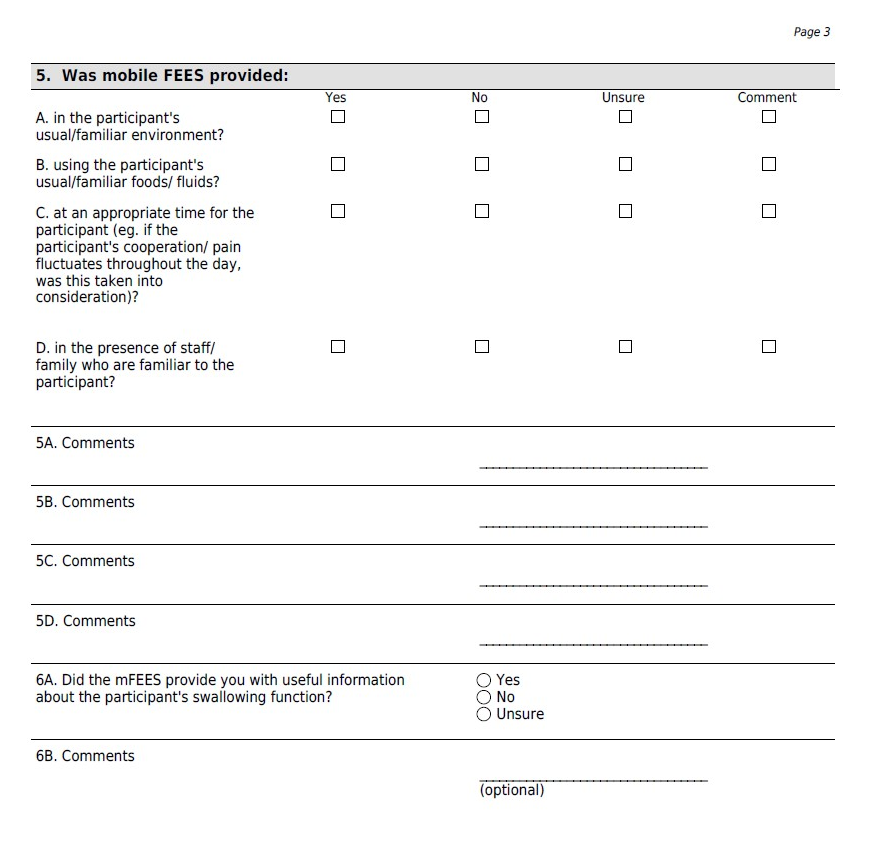

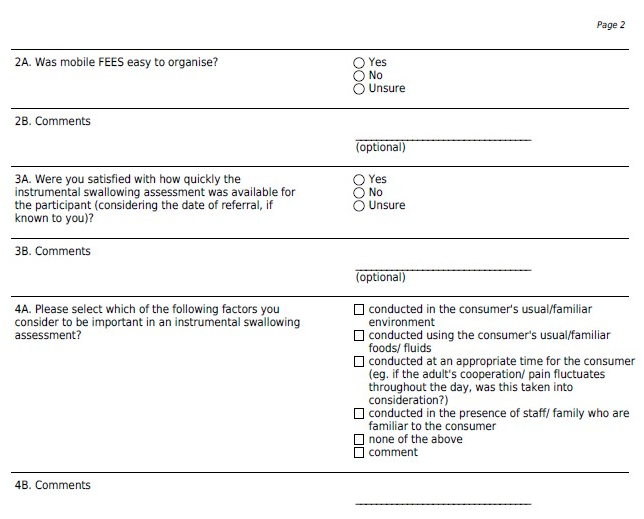

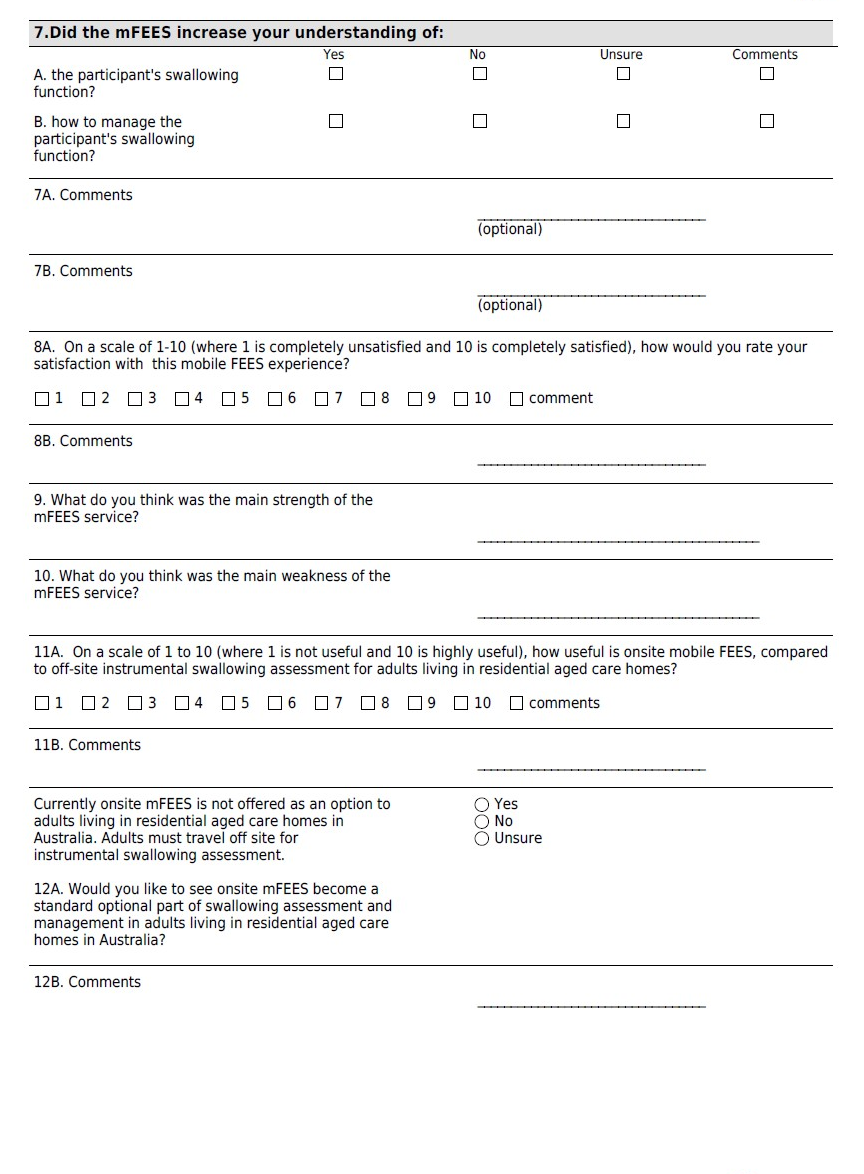

**Appendix 8.** Participant mFEES swallowing outcome measures

| **Resident Number** | **Oral Intake Trialled**  **(IDDSI classification)** | **The Yale Pharyngeal Severity Rating Scale Score** | | **Penetration-Aspiration Scale Score** |
| --- | --- | --- | --- | --- |
|  |  | Valleculae | Pyriform Fossae |  |
| 1 | L0 tsp | 2 | 2 | 1 |
|  | L0 sips | 3 | 3 | 1 |
|  | L2 tsp | 3 | 2 | 1 |
|  | L2 sips | 3 | 2 | 2 |
|  | L4 | 3 | 3 | 3 |
|  | L6 | 1 | 1 | 1 |
|  | L7 | 2 | 2 | 1 |
| 2 | L0 tsp | 2 | 2 | 3 |
|  | L0 sips | 2 | 2 | 6 |
|  | L4 | 2 | 2 | 2 |
|  | L6 | 1 | 1 | 2 |
|  | L7 | 1 | 1 | 1 |
| 3 | L0 tsp | 2 | 2 | 8 |
|  | L0 sips | 2 | 2 | 8 |
|  | L2 tsp | 3 | 2 | 3 |
|  | L2 sips | 3 | 2 | 3 |
|  | L4 | 3 | 3 | 3 |
|  | L6 | 4 | 3 | 3 |
|  | L7 | 4 | 2 | 2 |
| 4 | L0 tsp | 3 | 2 | 1 |
|  | L0 sips | 3 | 2 | 7 |
|  | L2 tsp | 3 | 3 | 1 |
|  | L2 sips | 3 | 3 | 8 |
|  | L4 | 3 | 2 | 1 |
|  | L6 | 3 | 2 | 1 |
|  | L7 | 3 | 1 | 1 |
| 5 | L0 tsp | 3 | 2 | 6 |
|  | L0 sips | 3 | 2 | 6 |
|  | L0 controlled swallow | 3 | 2 | 6 |
|  | L2 | 2 | 2 | 2 |
|  | L4 | 3 | 2 | 2 |
|  | L6 | 1 | 1 | 1 |
|  | L7 | 1 | 2 | 1 |
| 6 | L0 | 2 | 2 | 7 |
|  | L2 | 2 | 2 | 3 |
|  | L3 | 2 | 2 | 1 |
|  | L6 | 2 | 1 | 1 |
|  | L7 | 4 | 3 | 1 |
| 7 | L0 tsp | 1 | 1 | 1 |
|  | L0 sips | 1 | 1 | 8 |
|  | L0 tsp to clear pharyngeal residue | 1 | 1 | 6 |
|  | L2 tsp | 1 | 1 | 2 |
|  | L2 sips | 2 | 1 | 3 |
|  | L4 | 1 | 1 | 1 |
|  | L6 | 3 | 1 | 1 |
|  | L7 | 4 | 1 | 1 |
| 8 | L0 tsp | 2 | 2 | 1 |
|  | L0 sips | 2 | 2 | 8 |
|  | L0 sips with controlled swallow | 2 | 2 | 1 |
|  | L4 | 2 | 1 | 1 |
|  | L6 | 1 | 2 | 1 |
|  | L7 | 2 | 2 | 1 |
| 9 | L0 tsp | 2 | 2 | 2 |
|  | L0 sips | 3 | 3 | 3 |
|  | L2 tsp | 2 | 2 | 2 |
|  | L2 sips | 2 | 2 | 2 |
|  | L4 | 3 | 2 | 3 |
|  | L6 | 3 | 1 | 2 |
| 10 | L0 tsp | 2 | 2 | 1 |
|  | L0 sips | 2 | 2 | 1 |
|  | L2 | 3 | 3 | 1 |
|  | L4 | 3 | 2 | 1 |
| 11 | L0 tsp | 2 | 3 | 3 |
|  | L0 sips | 2 | 3 | 8 |
|  | L2 | 2 | 3 | 3 |
|  | L4 | 2 | 3 | 2 |
|  | L6 | 2 | 3 | 2 |
|  | L7 | 1 | 1 | 8 |
| 12 | L0 | 3 | 3 | 6 |
|  | L2 | 3 | 3 | 3 |
|  | L3 single sips | 3 | 3 | 3 |
|  | L3 liquid chaser for food residue | 3 | 3 | 8 |
|  | L5 | 5 | 1 | 8 |
